# Genomic Taxometric Analysis of Negative Emotionality and Major Depressive Disorder Highlights a Gradient of Genetic Differentiation across the Severity Spectrum

**DOI:** 10.1101/2025.01.30.25321336

**Authors:** Garrett W. Ennis, Travis T. Mallard, Javier de la Fuente, Camille M. Williams, Ted Schwaba, Elliot M. Tucker-Drob

## Abstract

A core question in both human genetics and medicine is whether clinical disorders represent extreme manifestations of continuous traits or categorically distinct entities with unique genetic etiologies. To address this question, we introduce *Genomic Taxometric Analysis of Continuous and Case-Control* data (GTACCC), a novel method for systematically evaluating continuity and differentiation of traits across the severity spectrum. GTACCC’s key innovation lies in binarizing continuous data at multiple severity thresholds, enabling the estimation of genetic continuity and differentiation within the trait and in its relation to other traits via multivariate models. We apply GTACCC to self-reported neuroticism data from UK Biobank (N= 414,448) and clinically ascertained major depressive disorder (MDD) data from the Psychiatric Genomics Consortium (ΣNeff = 111,221). We find that while neuroticism shares a considerable portion of its genetic etiology with MDD across the nonclinical, and even very low, range of (*r_g_* ∼ .50), genetic sharing increases monotonically across the severity spectrum, approaching unity only at the highest levels of severity (*r_g_* ∼ 1.0). Genomic structural equation models indicate that a single liability threshold model of negative emotionality is less consistent with the data than a multifactor model, suggesting that a gradient of genetic differentiation emerges across the spectrum of negative emotionality. Thus, within continuous measures of negative emotionality, partly distinct genetic liabilities exist at varying severity levels, with only the most severe levels associated with liabilities that approach equivalence to MDD genetics.

## Introduction

A fundamental question in human genetics and medicine is whether clinical disorders represent extreme manifestations of continuously distributed traits or categorically distinct phenotypes with distinct etiologies. This question is especially relevant to psychiatric disorders such as major depressive disorder, where symptoms can vary considerably outside the clinical range^1^. Discerning between continuous and categorical models of psychiatric illness is critical to a wide variety of scientific and clinical applications, including the design of empirical research (e.g., determining whether data can be aggregated across continuous and case-control studies), the development of nosological and diagnostic frameworks, and the appropriate application of therapeutics developed and validated for clinical populations to those with subclinical symptoms and vice versa. However, existing *taxometric* methods for empirically resolving this distinction rely on the analysis of phenotypic data^2–5^. Such approaches are particularly sensitive to the metric and distributional properties of the measures and fail to directly test for sharing versus differentiation of etiological factors across continuous and categorical phenotypes.

Here, we introduce *Genomic Taxometric Analysis of Continuous and Case-Control* data (GTACCC), a novel method for interrogating the typology of clinical disorders. GTACCC’s core innovation involves binarizing quantitative traits (e.g., continuous symptom measures) across multiple cut points, performing genome-wide association study (GWAS) analyses on each binarized phenotype, and then applying multivariate genetic methods to evaluate genetic continuity and differentiation across the severity spectrum. When appropriate external GWAS data are available, such as case-control data for the clinical diagnosis corresponding to the continuous symptom data, GTACCC also enables the analysis of genetic sharing between the external phenotype and the continuous phenotype across the severity spectrum. A particularly attractive attribute of GTACCC is that, by mapping the prevalence of individuals below each cut point to the cumulative normal distribution (Table S1), GTACCC is robust to departures from interval measurement of the quantitative trait that occur when, for example, an unbalanced set of mild and severe symptoms are queried. The quantitative trait must be measured in a population sample as opposed to a patient sample or a case-control sample in which cases have been oversampled (in contrast, the case-control GWAS of the clinical diagnosis may come from samples in which cases have been disproportionately ascertained). Additionally, the measure of the continuous trait must evince a wide enough range of scores to cover a meaningful span of severity. Careful consideration must also be given to whether the content measured by individual items or symptoms composing the continuous measures varies systematically with item or symptom severity; for optimal inference, symptom type should not be strongly related to symptom endorsement rates. In our empirical analysis, we conduct a range of sensitivity tests to assess these core assumptions.

We apply GTACCC to assess the continuity versus heterogeneity of the genetic etiology of negative emotionality across the severity spectrum, from normal-range neuroticism to clinically severe major depressive disorder (MDD). While it has been common in psychiatric genetics to treat continuous and case-control depression phenotypes as representative of the same liability^6,7^, some researchers have questioned this approach. Flint^8^, for example, has argued against the perspective that “MDD cases identified by minimal phenotyping are just less severe forms of MDD, and thus share the same genetic loci.” ^8^ He has proposed that “almost all GWAS have mapped a vulnerability to low mood or negative affect, a trait which is best termed dysphoria,” but has conceded that it is currently “too early to draw any definitive conclusions.” Using GTACCC, we find that although negative emotionality shares considerable genetic etiology with MDD across nonclinical and even very low levels (*r_g_* ∼ .50), genetic sharing increases monotonically across the severity spectrum, approaching unity (*r_g_* = 1.0) only at the most severe levels. Sensitivity tests reveal only a very weak tendency for more severe items to be more strongly related to MDD, indicating that the observed pattern is largely driven by the severity of the negative emotionality phenotype itself rather than by confounding of item content with severity.

## Results

Our analytic plan for primary analyses involving GTACCC of Neuroticism with respect to MDD was preregistered, and may be found at: https://osf.io/3wjyb/?view_only=75a8dc4249cc473885a78ae61f0a3b3d. Analyses of collateral phenotypes (e.g. subjective well-being, other psychiatric disorders, and other depression phenotypes) were not pre-registered.

### Overview of Method

GTACCC is an analytical framework that leverages GWAS data from a continuous trait or symptom set and, when available, one or more external traits (e.g., a corresponding case-control phenotype) to assess genetic continuity and differentiation along the severity spectrum. The continuous trait must be measured in a population-based and unascertained sample (it cannot be a case-only sample, a sample in which cases have been oversampled, or one that explicitly excludes cases).

The first step of GTACCC involves binarizing the continuous measure across a range of cut points corresponding to different severity levels, thereby creating a set of dichotomous variables, each with its own sample prevalence. In many circumstances, the number of cut points will be constrained by the resolution of the continuous measure (e.g. if the original variable is a sum of 12 dichotomous items, then only 12 cut points are possible). However, when the continuous variable is measured with high resolution, there is flexibility in the number and spacing of cut points, allowing a balance between precision and complexity (for example, it may not be necessary perform GWAS of each of 1000 cut points for a continuous measure of height). Each binarized version of the continuous trait is then used in genome-wide association analyses.

In the second step of GTACCC, the resulting GWAS summary statistics are entered into multivariate linkage disequilibrium score regression (LDSC) to estimate a genetic covariance or correlation matrix and its corresponding sampling covariance matrix, which represents the estimation errors of the elements in the genetic covariance or correlation matrix and their codependencies. The elements of interest (e.g., the genetic correlation between each binarized variable and the case-control phenotype) are then modelled as a function of their corresponding severity levels using a generalized least squares (GLS) estimator, using a weight matrix based on the corresponding elements of the sampling covariance matrix^9^. We avoid relying on the cut points themselves, which may suffer from lack of interval scaling. Rather, when the continuous trait does not have a desirable intrinsic metric, we map the prevalence of each constructed binary variable to the normal distribution and identify the corresponding locations along the normal distribution (i.e., a Z score relative to the latent distribution) as our severity index (Figure S1). The combined implementation of GLS and LDSC ensures that sample overlap resulting from the entry of different binarizations of the same continuous variable does not bias estimates, their standard errors, or *p* values.

In addition to formally modelling the magnitudes of genetic correlations as a function of severity using GLS, the GTACCC framework allows for the specification of a model within Genomic SEM, in which factors representing (partly) distinct dimensions of variation in genetic risk for different levels of severity are allowed to covary with one another and with the genetic component(s) of one or more external GWAS traits. As with GLS, the implementation of Genomic SEM using LDSC-derived matrices ensures that parameter estimates, standard errors, *p* values, and model fit statistics are not biased by sample overlap.

### GTACCC of Negative Emotionality in relation to Major Depressive Disorder

For a continuous measure of negative emotionality, we selected the 12-item Eysenck Neuroticism Questionnaire^10^ measured in 414,448 white British individuals in the UK Biobank. Aggregated neuroticism sum scores ranged from 0 to 12. We then binarized these neuroticism scores at 12 different cut points to create 12 binary variables, which were analyzed using GWAS with REGNIE to account for participant relatedness^11^ (Table 1). In the supplement, we provide results based on a subsample of unrelated participants in UK Biobank using PLINK (Table S2). Case-control GWAS summary statistics for clinically ascertained MDD (*N_cases_*= 45,591, *N_controls_* = 97,674) in European ancestry individuals were obtained from Wray et al. (2018)^12^, excluding data from both UK Biobank and 23andMe (where depression was measured using minimal or shallow phenotypes^13^). Effective sample sizes, mean *χ*^2^, and LDSC intercepts of the MDD and neuroticism summary statistics can be found in Tables 1. As expected, effective sample sizes and mean *χ*^2^ were largest at neuroticism cut points nearer to the center of the distribution, where prevalence rates of the binarized variables were closer to those of a balanced design^14^. LDSC intercepts indicated minimal, if any, inflation due to confounding.

**Table 1.** Descriptives of GWAS for each binarized Neuroticism variable.

| **GWAS** | **Cases/**<br>**Controls** | **Effective**<br>**Sample**<br>**Size** | **Mean**<br>**$\chi^2(1)$** | **LDSC**<br>**Intercept** | **Liability Scale**<br>**SNP $h^2$** |
| --- | --- | --- | --- | --- | --- |
| Neuroticism<br>Cut Point 1 | 354,588/<br>59,860 | 199,598 | 1.373 | 1.022 | 0.101<br>(0.003) |
| Neuroticism<br>Cut Point 2 | 314,430/<br>10,017 | 302,381 | 1.545 | 1.021 | 0.118<br>(0.002) |
| Neuroticism<br>Cut Point 3 | 268,427/<br>146,021 | 377,148 | 1.651 | 1.016 | 0.124<br>(0.002) |
| Neuroticism<br>Cut Point 4 | 223,092/<br>191,355 | 411,796 | 1.700 | 1.024 | 0.125<br>(0.002) |
| Neuroticism<br>Cut Point 5 | 180,528/<br>233,920 | 408,480 | 1.700 | 1.024 | 0.126<br>(0.002) |
| Neuroticism<br>Cut Point 6 | 141,718/<br>272,729 | 372,009 | 1.663 | 1.019 | 0.129<br>(0.002) |
| Neuroticism<br>Cut Point 7 | 107,004/<br>307,443 | 318,959 | 1.613 | 1.004 | 0.133<br>(0.002) |
| Neuroticism<br>Cut Point 8 | 78,905/<br>335,543 | 255,134 | 1.524 | 1.001 | 0.13<br>(0.002) |
| Neuroticism<br>Cut Point 9 | 54,540/<br>359,908 | 187,469 | 1.428 | 1.003 | 0.127<br>(0.003) |
| Neuroticism<br>Cut Point 10 | 35,522/<br>378,925 | 122,013 | 1.322 | 0.994 | 0.126<br>(0.003) |
| Neuroticism<br>Cut Point 11 | 20,506/<br>393,942 | 77,251 | 1.212 | 1.005 | 0.123<br>(0.005) |
| Neuroticism<br>Cut Point 12 | 8,977/<br>405,470 | 35, 669 | 1.114 | 1.018 | 0.085<br>(0.018) |

Note. The REGENIE-derived summary statistics of the neuroticism cut points estimated using data from the UK Biobank's white British ancestry subsample including related individuals ( $N = 414,448$ ) are reported. As described in Grotzinger et al. (2023), the sum of effective sample sizes is calculated by the equation $\sum v_k (1 - v_k) n_k$ , where $v_k$ is the sample prevalence for cohort $k$ , and $n$ is the sample size for cohort $k$ (note that UK Biobank is analysed as a single cohort, whereas meta-analyses are aggregated across multiple cohorts). The sum of effective sample sizes corresponds to an equivalently powered GWAS with balanced case/control proportions (50/50). In the column furthest to the right, we report the LDSC-estimated the genetic correlations between the GWAS in the expanded and reduced sample GWAS.
\* - For the neuroticism cut points, the population prevalence was set to be equal to the sample prevalence when estimating the liability scale heritability.

Genetic correlations between the binarized neuroticism variables ranged from ∼1.0 for adjacent cut points to ∼.8 for distal cut points (Figure S2), indicating some differentiation in the genetic etiology of neuroticism across the severity spectrum. The decay of genetic correlations with increasing distance of cut points from the initial cut point was statistically significant (*β*_l_ = −0.059, s.e. = 0.014, p = 4.00 e-05, which can be interpreted as −0.059 *r_g_* units per unit of standardized severity; Table S3). Note that the combined implementation of multivariate LDSC to estimate genetic correlations alongside dependencies in their estimation errors, and the use of GLS regression to estimate decay in the genetic correlations across cut points ensured that the estimated pattern was not driven by differences in the extent of shared estimation error across binary neuroticism GWAS.

Figure 1 depicts the trend of monotonically increasing genetic correlations between MDD and each of the binarized neuroticism variables as a function of neuroticism severity. For example, at the least severe level of neuroticism, the genetic correlation with MDD was estimated at 0.49 (s.e. = 0.076), while the estimated genetic correlation with MDD increased to 0.90 (s.e. = 0.13) at the most severe level of neuroticism. We conducted a GLS analysis in which we regressed the 12 genetic correlation estimates between each cut point and MDD on cut point severity (indexed in Z distribution units). Results indicated a significant linear (*β*_1_**=** 0.09, s.e. = 0.03, p = 0.0015), but not quadratic (*β*_1_ = 0.08 s.e. = 0.03, p = 0.006; *β*_2_ = 0.03, s.e. = 0.02, p = 0.121) association between cut point severity and the genetic correlations (Table S4).

**Figure 1.**
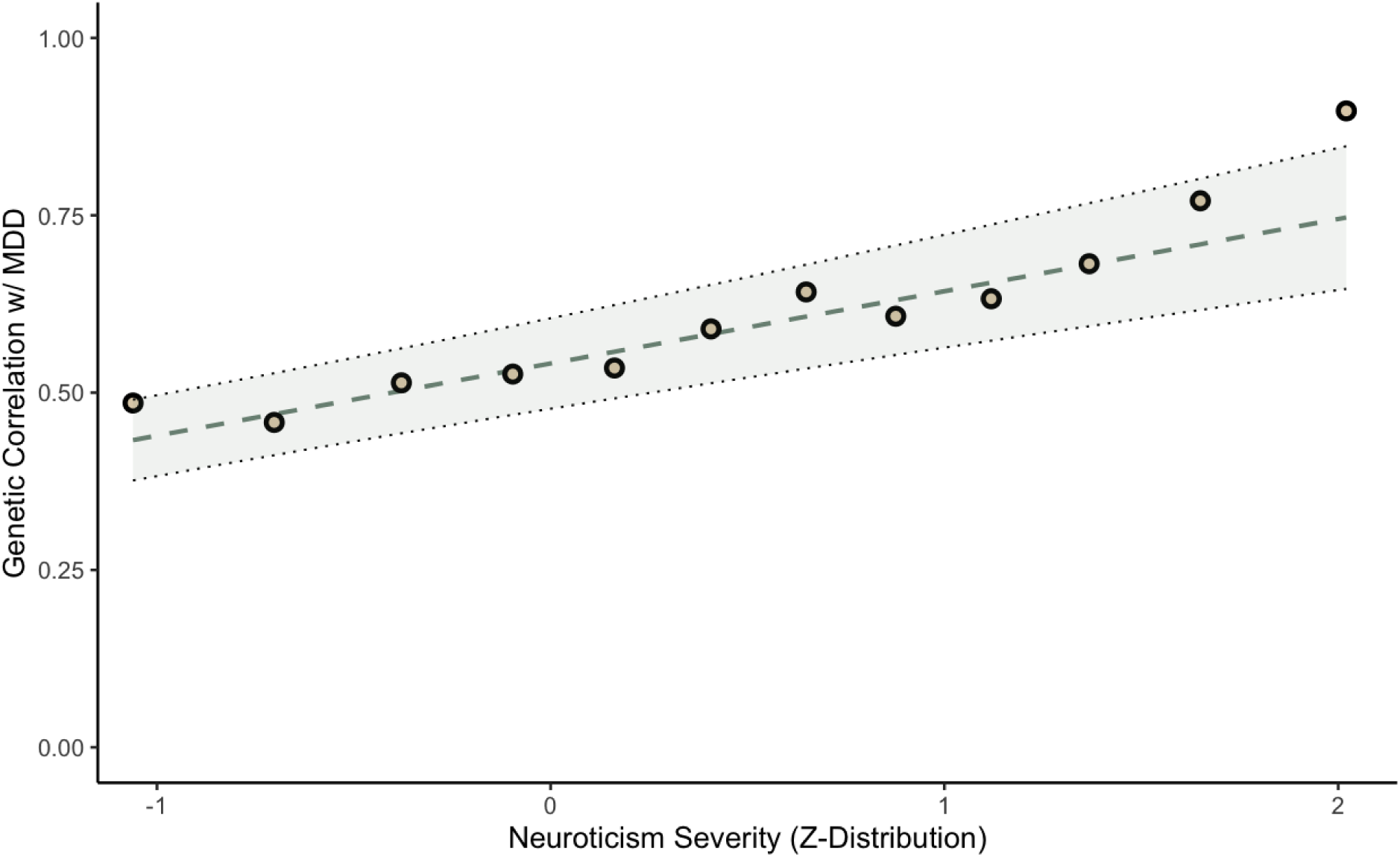
Genetic correlations between MDD and each neuroticism cut point. The line of best fit is shown, based on a linear regression model, with parameters estimated using generalized least squares (GLS). Shading around the line of best fit represents +/- 1 SE. GLS produces unbiased estimates of regression parameters and their standard errors when the independently and identically distributed (i.i.d.) assumptions required for OLS regression are not met, such that the dependent variables have potentially unequal and/or correlated estimation errors. GLS regression indicated that there is a statistically significant linear ( **=** 0.091, s.e. = 0.03, p = 0.0015), but not quadratic ( **=** 0.08, s.e. = 0.03, p = 0.006; = 0.034, s.e. = 0.022, p = 0.121) trend in the genetic correlations, indicating that as neuroticism severity increases, its underlying genetic etiology becomes more similar to MDD.

### Modelling of Severity Factors using Genomic SEM

We specified alternative factor models within Genomic SEM, allowing for one, two, and three severity factors, which were allowed to correlate with one another and with the genetic component of MDD. The two and three factor models (Figure 2) fit exceedingly well (two factor model: AIC = 270.174, CFI = 1, SRMR = 0.012; three factor model: AIC = 260.112, CFI = 1, SRMR = 0.012), compared to the one factor model (AIC = 1656.257, CFI = 0.998, SRMR = 0.053), thus indicating heterogeneity in genetic effects on neuroticism across severity levels. We found an increase in the genetic correlations between MDD and the severity factors across severity levels in both the two ( *rG_low,MDD_* = 0.45 (0.07), = 0.72 (0.08) and three ( = 0.45 (0.07), = 0.58 (0.04), = 0.72 (0.09) factor models.

**Figure 2.**
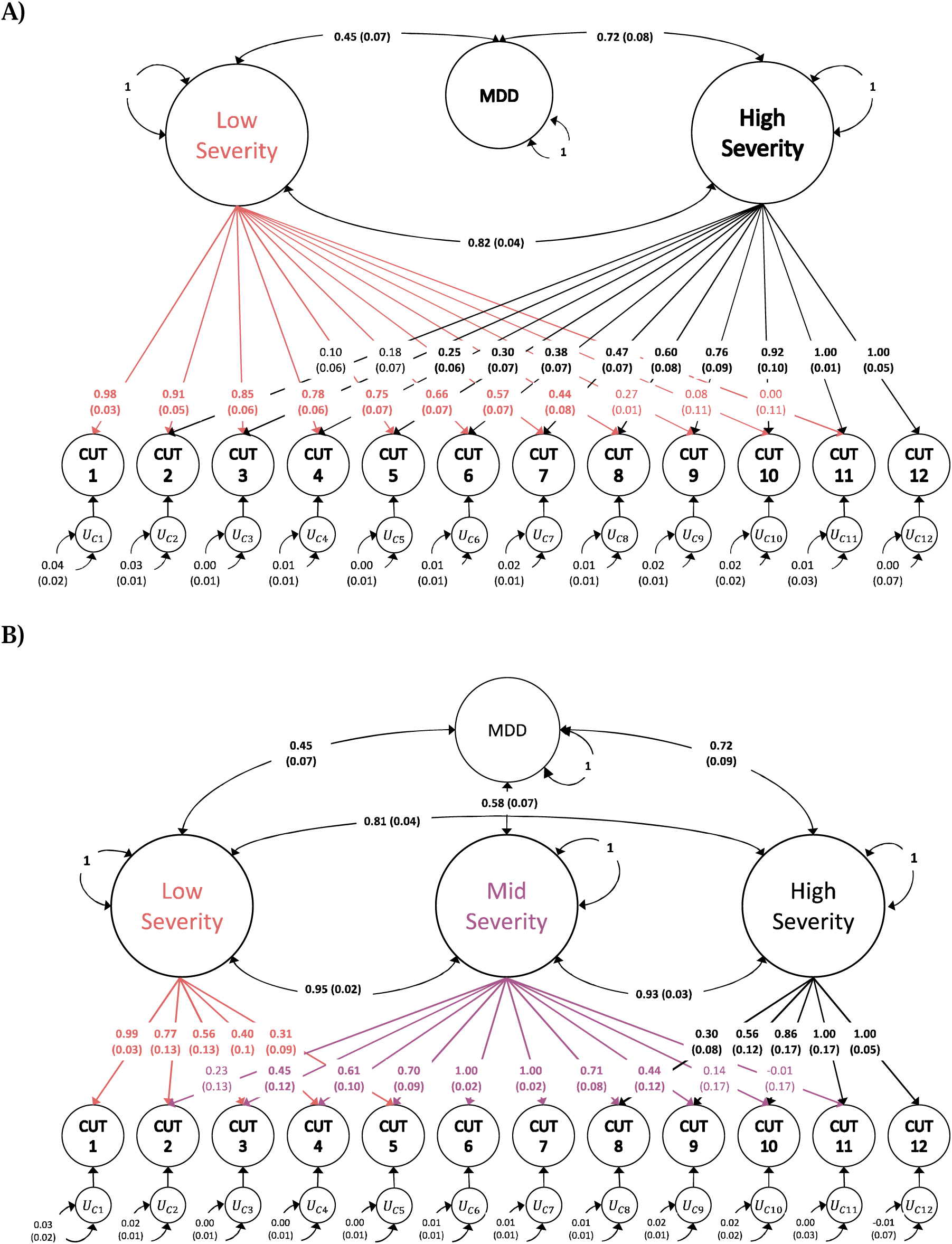
Standardized results (with standard errors) for **a)** two- and **b)** three-severity factor genomic structural equation models as estimated using GenomicSEM software. Circles represent inferred genetic variables. Covariance relationships among severity factors, and between the severity factors and MDD are represented by curved, double-headed arrows connecting variables. Variances for variables are represented by a curved arrow connecting the respective variable to itself. Both models fit the data extremely well, nearly identically (two factor model: AIC = 270.174, CFI = 1, SRMR = 0.012; three factor model: AIC = 260.112, CFI = 1, SRMR = 0.012), and far superior to a one factor model (AIC = 1656.257, CFI = 0.998, SRMR = 0.053).

Importantly, a gradient of loadings is readily apparent for the low and high severity factors with the highest loadings observed for the least (0.98, s.e. = 0.03) and most severe (1.00, s.e. = 0.05) cut points respectively. Progressing from least to most severe results in a gradual decrease in loadings on the low severity factor, and an increase in the loadings on the high severity factor. Additionally, within the three-severity factor model, the highest loadings for the middle severity factor are found in the middle of the range (1.00, s.e. = 0.02) and degrade as they move further from the center towards the extremes. While the severity factors evinced high intercorrelations correlations in both the two and three factor models, they are distinguishable from 1, indicating they index partly distinct genetic effects at different severity levels. Additionally, within the three factor model, the correlation between the low- and high severity factors (*rG_low,high_* = 0.81 (0.04) is markedly lower than the correlations between the low- and middle-severity factors (*rG_low,mid_* = 0.95 (0.02)), and the middle- and high-severity factors (*rG_mid,high_* = 0.93 (0.03)), indicating that the progressive gradient of genetic differentiation is also captured at the factor level.

### GTACCC of Negative Emotionality in relation to Continuous Neuroticism and Subjective Well-being

Given that our results revealed a gradient of genetic differentiation across different levels of neuroticism severity, a natural question that follows is what genetic effects are indexed when conducting a GWAS on a continuous neuroticism phenotype. We would expect that such a GWAS would index genetic signal most closely related to the intermediate levels of severity, i.e. that corresponding most closely to the cut point at the mean neuroticism sum score in the sample (UKB; M = 4.12, s.d. = 3.27).

We estimated the LDSC-derived genetic correlations between each level of neuroticism severity and those from the continuous non-binarized neuroticism phenotype. Results from these analyses reveal a high and flat pattern of genetic correlations (0.927 ≤ *r_g_* ≤ 0.991), with a slight inverse-U shape – the peak of which is centered approximately over a severity Z value of 0 (i.e., the value corresponding to the mean severity level in the sample; Figure S3). A GLS analysis (Table S4) indicated a significant quadratic (*β*_1_ = 0.001, s.e. = 0.011, p = 0.93; *β*_2_ = −0.025, s.e. = 0.01, p = 0.001), but not linear (*β*_1_ = −0.01, s.e. = 0.01, p = 0.37), trend in the genetic correlations, supporting this inference. These results indicate that the GWAS of the continuous trait most closely represents the genetic effects on that trait’s average severity within the analytic sample.

We specified the 3 Neuroticism severity factor model within Genomic SEM, as depicted in Figure 2B, replacing MDD with the continuous neuroticism GWAS phenotype. The model fit well (AIC = 147.38, CFI = 1, SRMR = 0.004). Consistent with the GLS analyses, the middle severity factor was more highly correlated with continuous neuroticism (*rG_mid,neu_* = 0.99, s.e. = 0.02) than were the low (*rG_low,neu_* = 0.96, s.e. = 0.03) and high (*rG_high,neu_* = 0.92, 0.03) severity factors.

We additionally conducted a GTACCC analysis of neuroticism in relation to a GWAS meta-analysis of subjective well-being based primarily on continuous measures of well-being (Supplementary Information; Figure S4; Table S4). As expected, we observed no discernable association between neuroticism severity and its association with subjective well-being. In contrast, had the subjective well-being GWAS been based on a binary phenotype, in which case status was based on extremely high levels of subjective well-being, we would have expected the strongest genetic correlations with Neuroticism to occur at the lowest levels of neuroticism severity, as very low levels of neuroticism would be expected to correspond more closely to very high levels of subjective well-being.

### Gradient of Genetic Differentiation is not an Artifact of Sample Prevalence

In contrast to observed scale heritability estimates, which depend on the sample and population prevalences of the binary trait, genetic correlation estimates are mathematically expected to be invariant across sample and population prevalences. Nevertheless, to empirically confirm that the observed pattern of increasing genetic correlations between neuroticism and MDD across severity levels was not an artifact of variation in prevalence rates across the binarized neuroticism phenotypes, we down-sampled to obtain balanced case/control proportions (50%) in both low and high severity cut points. To avoid complexity associated with down sampling data in the context of family structure, we used GWAS summary statistics for unrelated individuals, estimated in PLINK. For the lower severity cut point, we selected the 1^st^ cut point (n = 81,886), and for the higher severity cut point, we used the 11^th^ cut point out of 12 (n = 27,560), as down sampling to 50% case prevalence for the 12^th^ cut point would have reduced sample too drastically (n = 12,096) to accurately estimate genetic correlations using LD Score Regression. Using these balanced prevalence GWAS phenotypes, genetic correlations with MDD were estimated to be 0.51 (se = 0.08) for cut point 1 and 1.03 (se = 0.31) for cut point 11. As expected, the standard errors were considerably larger when estimating genetic correlations using these reduced samples. Nevertheless, the comparison of point estimates suggests that the differences in the genetic sharing between neuroticism and MDD across cut points persists after controlling for differences in case prevalence.

### Item-level Genetic Correlations with MDD Across Item Severities

We conducted analysis the genetic correlation between individual neuroticism items and MDD as a function of item severity, as indexed by item endorsement rates (items with lower endorsement rates index rarer symptoms and are thus inferred to be more severe). We found only a very weak tendency for more severe items to be more strongly related to MDD (Figure S5), indicating that the observed pattern of association between neuroticism severity and MDD is largely driven by the severity of the negative emotionality phenotype itself rather than being driven by specific items with low endorsement rates being more strongly genetically correlated with MDD.

### GTACCC of Negative Emotionality in relation to 13 Psychiatric Disorders and their Transdiagnostic Factors

As an exploratory analysis, we sought to determine whether the pattern of increasing genetic associations between negative emotionality and psychiatric disorders as a function of severity level generalized beyond MDD. To do so, we estimated the LDSC-derived genetic correlations between the 12 neuroticism severity cut points and 13 psychiatric disorders corresponding to five domains (Internalizing Disorders, Thought Disorders, Neurodevelopmental Disorders, Compulsive Disorders, and Substance Use Disorders), as well as 5 broad factors of transdiagnostic risk corresponding to each of these domains. Disorders varied considerably from one another in their overall magnitude of genetic correlation with negative emotionality, but we observed an overall pattern of increasing genetic correlations across severity levels (Table S7; Figure 3; Figures S9-13). Of the 13 individual disorders, the pattern of increasing correlation with negative emotionality at higher severity levels was significant for all but Anorexia Nervosa, Tourette’s Syndrome, Anxiety Disorders (for which standard errors were especially large), and Cannabis Use Disorder. At the level of the factors (Figure 3), this association was significant for all domains (Internalizing Disorders, Thought Disorders, Neurodevelopmental Disorders, and Substance Use Disorders) but the Compulsive Disorders domain (Table S7).

**Figure 3.**
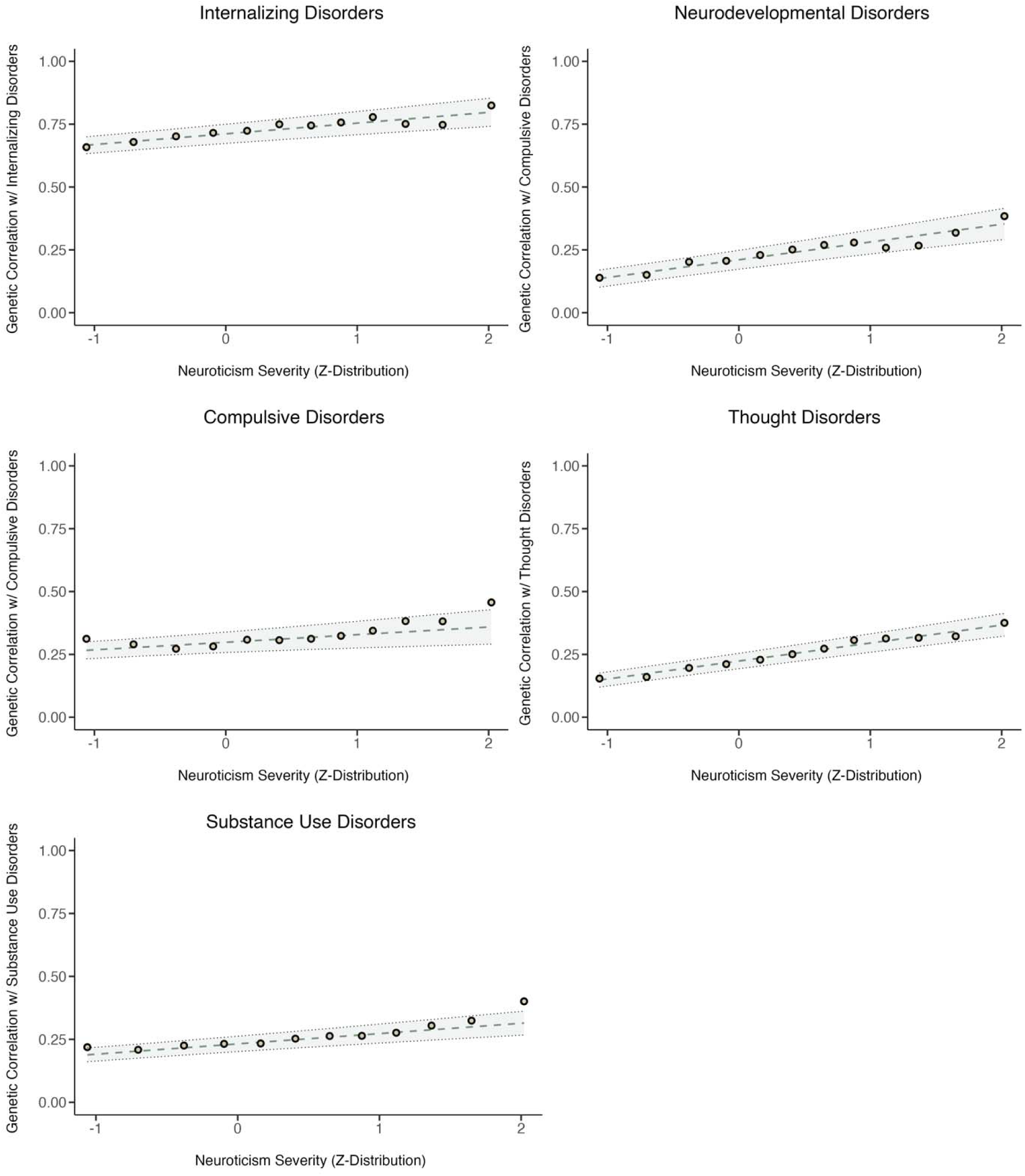
Genetic correlations between 5 transdiagnostic factors of psychiatric risk and each neuroticism cut point. The line of best fit is shown with parameters estimated using Generalized Least Squares (GLS) regression. GLS regression indicated that there were significant linear trends in the genetic correlations between neuroticism severity and 4 factors: Thought (*β*_1_ = 0.071, s.e. = 0.012, p = 1.46e-09); Neurodevelopmental (*β*_1_ = 0.071, s.e. = 0.018, p = 1.11e-04); Internalizing (*β*_1_ = 0.042, s.e. = 0.013, p = 0.002); and Substance Use (*β*_1_ = 0.041, s.e. = 0.013, p = 0.002), but not for the Compulsive factor (*β*_1_ = 0.03, s.e. = 0.019, p = 0.121). Complete GLS estimates are reported in Table S7.

### GTACCC of Negative Emotionality in relation to Alternative Binary Depression Phenotypes

We have reported results of GTACCC of Neuroticism in relation to MDD using our pre-registered primary outcome, which is a GWAS meta-analysis of case-control studies of clinical ascertained MDD from Wray et al., (2018)^12^, specifically excluding self-report measures and minimal phenotypes. As an exploratory analyses, we sought to gauge the extent to which the pattern of results obtained using the primary (clinically ascertained) MDD outcome would also be observed when using alternative depression phenotypes. To do so, we obtained GWAS summary statistics for eight different binary measures of depression (Table S9) derived from the UK Biobank by Cai et al (2020).^13^ All eight of these phenotypes, except for the electronic medical records, were constructed using self-reports (of symptoms, help seeking, and/or a depression diagnosis), using criteria varying in their levels of mapping to clinical diagnostic criteria, such that some are considered “minimal phenotypes” and other are considered “strictly defined.” We estimated the genetic correlations between each of these 8 phenotypes and the 12 neuroticism cut points, which we submitted to GLS regression (Figure S14; Table S9).

Overall, genetic correlations with neuroticism were moderate to high and we observed similar patterns of increasing genetic sharing between negative emotionality and depression at higher severity levels (Figure S14; Table S9), with all but two of the alternative depression phenotypes (LifetimeMDD and MDDRecurr, for which sample sizes were among the smallest and SEs among the largest) reaching significance at p < 0.05. This pattern was generally similar to that obtained for our primary MDD outcome, and for other the internalizing disorders (Figure S12, Table S7) and the broader Internalizing factor (Figure 3; Table S7). We did not observe stark differences in patterns according to whether Cai et al. (2020) designated the depression phenotype as minimal vs. strict, but we did observe that the linear relation between severity and genetic correlations were steepest when case status was based primarily on having seen a doctor for nerves, anxiety, tension, or depression, or was based electronic medical records (.062-.105 *r_g_* units per SD increase in severity), and shallower when case status was based primarily on self-reported symptoms (.026-.043 *r_g_* units per SD increase in severity).

## Discussion

Distinguishing between continuous and categorical disease models of clinical disorders is critical for nosology, therapeutic development, and clinical practice. This issue is especially relevant to psychiatric disorders. The diagnostic criteria for MDD constitute disruptive levels of negative emotion (with low mood and anhedonia being the cardinal symptoms) lasting over a period of at least two weeks that produce clinically significant distress or impairment in social, occupational, or everyday functioning^15^. However, negative emotionality varies considerably outside of the clinical range in the general population, and it has been unclear whether the genetic etiology of such variation is distinct from that which distinguishes clinically severe MDD cases from unaffected individuals in the general population. In this study, we introduced *Genomic Taxometric Analysis of Continuous and Case Control* data (GTACCC), a method for distinguishing between continuous and categorical models of genetic architecture. In our analysis of negative emotionality and MDD, we find evidence for a gradient of genetic differentiation across the spectrum of severity of negative emotionality such that while there is a portion of the genetic architecture which is common across all levels of severity, the genetic effects distinguishing individuals at specific levels of severity become progressively more differentiated the further apart those severity levels are from each other. Only at the most severe levels of negative emotionality do genetic effects strongly converge with those on MDD, indicating that the distinction between genetic etiologies of neuroticism and MDD may not be a matter of type but of degree.

Previous attempts to determine whether there exists a substantive, underlying categorical difference between individuals with clinically severe and sub-clinical levels of depressive symptoms have been phenotypically, rather than genetically, based and have produced equivocal results^16,17^. This may be due to having relied heavily on characteristics of the joint distribution of items or symptom measures in a sample, which may be affected by factors such as sample composition or the psychometric characteristics of the measures used (such as when severe symptoms are disproportionately queried)^18,19^. In contrast, by mapping the prevalence of individuals below each cut point to the cumulative normal distribution, GTACCC is robust to departures of the continuous measures from interval measurement that occur when, for example, an unbalanced set of mild and severe symptoms are queried. Moreover, whereas previous approaches have often lacked inferential statistics, instead relying on a subjective interpretation of patterns of relationships within the data^4,20^, GTACCC employs a GLS estimator in combination with multivariate LDSC estimator to model differentiation of genetic effects as a function of trait severity, producing parametric estimates, confidence intervals, and *p* values indexing the extent of such differentiation. Finally, in contrast to phenotypic approaches, which are agnostic to etiology, GTACCC directly models sharing and differentiation of *genetic etiology* across continuous and case-control phenotypes^1–4^.

Our findings suggest that hidden within continuous measures of negative emotionality are genetic effects specific to different levels of severity – such that there is partly a common genetic basis to phenotypic variation that is shared across all severity levels, alongside a continuous gradient of genetic differentiation across severity levels. Factor models were specified within Genomic SEM to test for the presence of distinct liabilities which could be used to differentiate the genetic component of variation in negative emotionality in relation to clinical MDD (Figure 2). We found that both the two- and three-severity factor models fit far better than a single common factor model, providing further evidence of a gradient in the genetic component of negative emotionality. Consistent with the inference that genetic etiology differentiates relatively continuously across the severity spectrum, the three-factor model indicated stronger genetic correlations among adjacent severity factors (low vs. mid, and mid vs. high) than among distal factors (high vs. low). Nevertheless, all factors were moderately to strongly genetically correlated, indicating that there is also a sizable amount of shared genetic etiology across different levels of negative emotionality.

Our findings are only partially consistent with dimensional theories of psychopathology, such as the HiTOP (Hierarchical Taxonomy of Psychopathology) model^21,22^, which treat clinical disorders as simply constituting extremes of continuously distributed dimensions. They are also only partially consistent with categorical models of psychopathology, which treat clinical diagnoses as entirely distinct from variability in the subclinical and normative regions of the severity distribution. We observed only a very weak tendency for more severe items to be more strongly related to MDD, indicating that the observed pattern is largely driven by the severity of the negative emotionality phenotype itself rather than by specific items with low endorsement rates and stronger genetic overlap with MDD. Importantly, however, individuals who endorsed more neuroticism items (those reporting greater neuroticism severity) were necessarily more likely to have endorsed a wider breadth of facets of neuroticism (e.g. mood lability, interpersonal distress, and anxious worry; Figure S8). This may indicate that the genetically distinct components of MDD relate to risk for the co-occurrence of multiple aspects of negative emotionality. It will be valuable for ongoing investigations motivated by both dimensional and categorical models to deploy multivariate methods for further elucidating these shared and unique genetic effects.

We found that the pattern of increasing genetic sharing between negative emotionality and psychiatric disorders as a function of severity of negative emotionality generalized across a wide range of disorders beyond MDD or even the internalizing domain of psychopathology. Several substance use disorders, neurodevelopmental disorders, and thought disorders also exhibited this pattern, even when their overall magnitude of genetic association with negative emotionality was itself only moderate. While negative emotionality has itself been implicated as a core risk factor for a wide range of psychiatric disorders, this finding indicates that severe negative emotionality may more specifically drive such transdiagnostic risk. Moreover, when using alternative binary depression phenotypes, we observed steeper linear relation between severity of negative emotionality and its association with depression when case status was based primarily on having seen a doctor for nerves, anxiety, tension, or depression, or was based electronic medical records (.062-.105 *r_g_* units per SD increase in severity), and shallower when case status was based primarily on self-reported symptoms (.026-.043 *r_g_* units per SD increase in severity). This may indicate that a key feature distinguishing the genetic etiology of severe depression from normal range negative emotionality is not the constellation of specific symptoms per se, but the that they are distressing enough to warrant engagement with the healthcare system.

The GTACCC approach that we have introduced here may be of considerable utility for discerning continuous from categorical models of disease across a wide range of phenotypes. Importantly, care must be taken, as we did here, to ensure that the data meet certain requirements to support the inferences made. First, the continuous trait must be measured in a population sample as opposed to a patient sample or a case-control sample in which cases have been oversampled. Second, the resolution of the continuous measure must be sufficient to estimate GWAS across a range of severity levels. Measures that are too coarse may obscure a gradient of genetic differentiation when pooling individuals into too broad of categories. Similarly, if the measure used exhibits dramatic floor or ceiling effects, this will substantially limit the observable range of severity. Third, careful consideration must be given to whether the content measured by individual items or symptoms composing the continuous measures varies systematically with item or symptom severity. Finally, when multiple traits (e.g., a continuous and a case-control trait) are examined together within GTACCC, there must be sufficient rationale examining them in relation to one another.

It is important to consider the limitations of the current study. First, we examined a continuous measure of neuroticism in relation to clinical MDD. It may have been preferable to employ a continuous measure of MDD symptomology, rather than neuroticism. However, we are unaware of any large-scale symptom level data for MDD in a general population sample (e.g. in UK biobank, many depression symptoms are only queried if gating symptoms are endorsed)^23^, and it is possible that if clinical symptom inventories were used in such samples, dramatic floor effects would limit their utility. That we obtained an estimate of genetic correlation between neuroticism and MDD approaching 1.0 at the highest level of neuroticism severity is perhaps even more impressive given that neuroticism is not itself a direct inventory of MDD symptomology. Second, the neuroticism measure itself was composed of dichotomous (yes or no) items. Had more continuous item-level information been available (e.g., by allowing for responses on a 7-point Likert scale), further opportunities to test differentiation of genetic etiology across severity levels would have been possible at the level of individual items or symptoms. Finally, we analyzed neuroticism as a unidimensional construct. Analyzing specific facets of neuroticism would have allowed for more complex applications of the GTACCC framework than those described here.

In conclusion, we have introduced the GTACCC framework for testing the extent to which clinical disorders represent extreme manifestations of continuously distributed traits or categorically distinct entities with qualitatively distinct genetic etiologies. In our application of GTACCC to negative emotionality and clinical MDD, we find evidence for a gradient of genetic differentiation across severity levels of negative emotionality. The genetic etiologies of neuroticism and MDD overlap considerably even at the very lowest levels of severity (*r_g_* ∼ .50). However, at increasing severity levels, genetic sharing increases monotonically, approaching unity (*r_g_* ∼ 1.0) only at its very severe levels. Factor models specified within Genomic SEM indicate that a single liability threshold model of negative emotionality is less consistent with the data than a multifactor model. Thus, hidden within continuous measures of negative emotionality are genetic effects specific to different severity levels, with only the most severe levels approaching equivalence with the genetic etiology of MDD. These results are partly consistent with both the continuous and disease models of negative emotionality and MDD. GTACCC provides considerable opportunities for testing continuity and differentiation of genetic etiology for dimensions and disorders not considered here. To aid researchers in conducting their own applications of GTACCC, we provide tools for conducting GTACCC within the free, open-source Genomic SEM environment.

## Methods

### Genomic Taxometric Analysis of Continuous and Case Control Data

GTACCC is an analytical approach that leverages genetic data for a continuous trait or symptom set and, when available, one or more external traits (e.g., a corresponding case-control trait) to assess genetic continuity and differentiation along the severity spectrum. First, a set of GWASs are conducted on the continuous trait of interest by progressively binarizing along different levels of severity to estimate the SNP effect which distinguish individuals above and below the given cut point. The resulting GWAS summary statistics are then input into multivariate LD Score Regression to estimate a matrix of genetic covariances (S) or genetic correlations (R) between severity levels and, when available, between severity levels and one or more external traits of interest, along with an associated matrix of sampling covariances (V_S_ or V_R_) that index estimation errors and their codependences across cells of S or R. The genetic covariance or genetic correlation matrix is then subset to a vector of elements of interest, and the corresponding sampling covariance matrix is subset accordingly. These matrices are then submitted to multivariate analysis using generalized least squares (GLS) and/or Genomic SEM. For the primary analyses reported for this paper, we model elements drawn from the standardized S matrix, and its corresponding sampling covariance matrix. Although the standardized S matrix can be considered equivalent to a genetic correlation matrix, the behavior of its multivariate sampling distribution may differ from that associated with a more direct estimate of the genetic correlation matrix (R). In the supplemental note, we explicate this difference, and we report results from analyses based on elements drawn from R and V_R_, which were very similar those of the primary analyses reported in the main text.

### Generalized Least Squares

GLS is similar to weighted least squares (WLS) regression in that the fit function is weighted by the precision of the individual estimates in y^9,24^. However, GLS further expands on WLS by taking into account the dependencies in estimation errors in the y estimates, which can result from sample overlap (such as when the outcomes represent genetic correlations among phenotypes formed by binarizing the same continuous measure at different cut points). The vector of GLS coefficients, including the regression intercept, is estimated as follows:

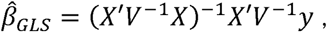

where X is the predictor set (including a vector of 1s for a regression intercept), y is the vectorized (sub)set of elements within S being modelled, and V is the sampling covariance matrix of y. The corresponding covariance matrix of the GLS coefficients is estimated as:

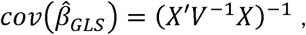

where the square root of the diagonal of 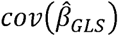 are the standard errors of the GLS coefficients^9,24^. We provide functions to subset the relevant components of the LDSC-derived S and V matrices and performing GLS based on these summary data. GLS permits the regression of genetic correlations of interest (as the y variable) as a function severity levels (as the X variable) while accounting for the estimation errors of the genetic correlations and dependencies in those estimation errors (represented in V). When there is an intrinsic metric of the continuous measure, it may be sensible to use the cut point value itself as the index of severity. However, when the metric of the continuous measure is arbitrary, we advise that the observed prevalence for each cut point be mapped to a location along a normal distribution (e.g. using the qnorm() function within R) to assign it a severity level on a standardized scale.

### GTACCC Models within Genomic SEM

In addition, or as an alternative to formally modelling the patterning of genetic correlations as a function of severity using linear or polynomial GLS, a GTACCC-based Genomic SEM model can be specified to reflect factors representing (imperfectly) correlated dimensions of variation in genetic risk at different regions of the severity spectrum.

Each of the k binarized versions of the continuous GWAS trait are modelled as reflecting m common factors and k unique genetic factors according to:

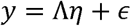

Where y is a k x 1 vector of indicators, *η* is an m x 1 vector of correlated latent variables (i.e. factors), Λ is a k x m matrix of factor loadings connecting the set of indicators to the latent variables, and *ε* is a k x 1 vector of residuals representing unique genetic factors (residuals) specified to be independent^25^. This factor model can be expanded to include genetic associations between each latent and one or more external GWAS traits, such as clinical MDD. We would generally expect the unique genetic factors (*∊*) to contribute trivially to each individual binarized trait, but the number of m latent factors needed to explain the pattern of genetic variation and covariation among the binary factors is unknown.

In one extreme, in which the liability threshold model perfectly holds, only a single common factor is required as the assumption under this model is that the same genetic effects underly variation across all points along the severity spectrum. However, more relaxed models may be required to describe variation the genetic effects on a given trait. Such models may specify multiple (correlated) common factors, with each factor reflecting shared genetic variation relevant to different levels or ranges of disease severity. For instance, a two-factor model can be specified in which the lowest severity GWAS phenotype (the constructed binary variable based on the lowest cut point) only loads on the first factor, such that the factor represents genetic liability to low severity, and the highest severity GWAS phenotype (the constructed binary variable based on the highest cut point) only loads on the second factor such that it represents genetic liability to high severity. All other GWAS phenotypes load on both factors and the two factors are allowed to correlate with one another. In this case, the anchoring of the cut points related to the most extreme levels of severity would estimate how genetic variation at the lowest and highest level explains variation along the range of cut points which compose the factors. Additionally, the gradient of loadings within each factor quantifies the extent to which the severity-specific genetic factors explains variability in symptoms at the more central levels of severity. A similar approach can be taken to specify a three-factor model corresponding to genetic liability to low, mid, and high levels of severity. Factor models can be empirically compared using model fit indices, such as SRMR, CFI, and AIC as detailed in Grotzinger et al. (2019)^25^.

In the present application, we specified three different severity factor models corresponding to one, two and three severity factors. In the one factor model, a single common factor was specified to underly genetic variation in all cut points. In the two-factor model (Figure 2A), a low severity factor was specified to underly genetic variation in cut points 1-11, and a high severity factor was specified to underly genetic variation in cut points 2-12. In the three-factor model (Figure 2B), the low severity factor was specified to underly genetic variation in cut points 1-5, the middle severity factor was specified to underly genetic variation in cut points 2-10, and the high severity factor was specified to underly genetic variation in cut points 8-12.

### Sources of GWAS Data

#### Major Depressive Disorder (MDD)

We used GWAS summary statistics of MDD for European ancestry individuals from Wray et al., (2018)^12^, which was a large-scale collaboration from the Psychiatric Genetics Consortium (PGC) MDD Working Group. We opted to use the summary statistics provided that excluded 23andMe and UK Biobank (UKB) to obtain a case-control MDD phenotype composed specifically of clinically defined cases. Within the remaining datasets, cases were determined either by structured clinical interview or electronic health record data, whereas in the 23andMe and UKB datasets, depression cases were partially determined using minimal phenotypes or self-report depression diagnosis. This reduced set of summary statistics was composed of 29 case-control samples from the PGC29 cohort as well as 4 additional cohorts from the deCode (Ripke et al., 2013), GenScotland^26^, GERA^27^ and iPSYCH^28^ altogether composing a total of, 45,591 cases and 97,674 controls, with a sum of effective sample sizes^29^ of 111,221.

### Continuous Measure of Negative Emotionality

Negative emotionality was indexed by a 12-item measure of neuroticism in UK Biobank (UKB). The UKB dataset is a population-based cohort with ∼500k individuals from both rural and urban communities in the UK^30^. At recruitment, individuals answered questionnaires detailing their socio-economic, demographic, lifestyle, and health-related factors, and provided blood, urine, and saliva samples for genetic, proteomic and metabonomic analysis^31^. Data was collected between 2006 and 2010, and at that time participants were between 39 to 73 years old. Our analyses were restricted to self-reported white British individuals to avoid confounding due to large scale population stratification.

Neuroticism is a personality trait conceptualized as an individual’s level of negative emotionality defined by the frequency, duration, and intensity in their tendency towards negative emotionality^32,33^. Neuroticism indexes low mood, stress sensitivity, lack of emotional control, and emotional lability. Previous work has indicated that neuroticism is highly phenotypically^34–36^ and genetically^7,37–40^ correlated with depression (*r_g_*(Neu, MDD) ∼ 0.62-0.68, *r_g_*(Neu, Depressive Symptoms) ∼ 0.75 – 0.82).

Items were drawn from Eysenck, Eysenck & Barrett^10^ (1985;Table S1). Participants could respond to these questions in four ways: “Yes” (scored as 1), “No” (scored as 0), “Do not know” (scored as missing) and “Prefer not to answer” (scored as missing). Following Nagel et al., (2018)^38^, individuals were dropped from our analyses who completed <9 neuroticism items, indicating that they provided answers of “I don’t know” or “Prefer not to answer” to more than 3 items on the Eysenck neuroticism Questionnaire. Scores across all non-missing items were averaged for each person and multiplied by 12 producing scores ranging from 0 to 12. This continuous measure was then binarized at integer cut points ranging from 1 to 12 (scores below cut point = 0, scores at or above cut point =1).

To control for confounding due to population stratification, GWAS of binarized neuroticism phenotypes were restricted to self-reported white, British individuals within the UKB who were missing less than 2% of their genotype data (*n* = 414,448). We conducted GWAS using REGNIE ^11^ which utilizes a whole genome, machine learning regression method capable of controlling for small scale population structure, allowing for the inclusion of related individuals. We also conducted sensitivity analyses using GWAS of binarized neuroticism phenotypes in a subset of unrelated individuals using PLINK. Results from these GWAS were very similar to those obtained from REGENIE (Table S2-4. Our pipeline involved applying LDSC to GWAS summary statistics. Thus, because LDSC only employs common Hapmap3 variants^41^, GWAS were performed only on those SNPs. Further quality control was implemented to limit SNPs to those with MAF > 0.01, MAC > 100, excluding those with missing genotype rate of >10%, and HWE < 10^-l5^, leading to a final SNP count of 1,033,032.

## Acknowledgements

This research was supported by National Institutes of Health (NIH) grant R01MH120219. EMTD is member of the Population Research Center (PRC) and the Center on Aging and Population Sciences (CAPS) at The University of Texas at Austin, which are supported by NIH grants P2CHD042849 and P30AG066614, respectively. We are grateful to Lea Davis for helpful feedback on previous drafts of this article.

## Data and Code Availability

Summary statistics for the GWAS of neuroticism at each cut point in both PLINK and REGENIE will be made available at https://osf.io/qc4ue/

Code for key analyses reported in this paper may be found at https://osf.io/8j2fx/?view_only=4e90349d3ded46d7a7983af3add081a3

GenomicSEM software has been updated to include new functions to subset genetic covariance matrices and fit generalized least squares models in support of GTACCC. Genomic SEM is freely available at https://github.com/GenomicSEM/GenomicSEM

Tutorials for implementing these new functions can be found on the GenomicSEM wiki at https://github.com/GenomicSEM/GenomicSEM/wiki/8.-Tutorials

