## Supplementary Methods and Figures for "Genomic Taxometric Analysis of Negative Emotionality and Major Depressive Disorder Highlights a Gradient of Genetic Differentiation across the Severity Spectrum"

#### **This PDF file includes:**

Supplementary Methods:

*Genetic correlations with continuous measures of neuroticism*

*GTACC in relation to a Continuous GWAS of Subjective Well-being*

*Item-level Genetic Correlations with MDD Across Item Severities*

#### Re-Estimating GLS Models Using Direct Estimates of Genetic Correlation

Supplementary Figures

**Other supplementary materials for this manuscript found in the Supplementary Tables document:**

Supplementary Methods

***Genetic correlations with continuous measures of neuroticism***

We repeated our primary analyses, in which we estimated genetic correlations between each of the 12 binarized neuroticism GWAS and a GWAS of the continuous measure of neuroticism from UK Biobank (<http://www.nealelab.is/uk-biobank/>). Results are presented in Figure S3 and Table S4. Genetic correlations ranged from 0.91 (at cut point 11) to 0.99 (at cut points 4 and 5). A GLS regression analysis of the relation between cut point severity and genetic correlations with continuous neuroticism indicated an inverted u-shape with a significant quadratic trend ($\beta_{1}$ = 0.001, s.e. = 0.01, P = 0.934; $\beta_{2}$ = -0.02, s.e. = 0.01, P = 0.0013), but nonsignificant linear trend $\beta_{1}$ = -0.021, s.e. = 0.02, P = 0.0176). Of note, the average neuroticism sum score within the UK Biobank is 4.12, lying between the values of the two cut points with the greatest genetic correlations with a continuous measure of neuroticism. This indicates that the GWAS of the continuous trait represents the genetic architecture of that trait’s average severity within the analytic sample in which the GWAS is most well powered.

***GTACC in relation to a Continuous GWAS of Subjective Well-being***

We repeated our primary analyses, substituting the summary statistics from a metanalytic GWAS of subjective well-being from Okbay et al., 2016^1^ (excluding 23andMe) in place of MDD. Subjective well-being has been consistently found to evince strong genetic correlations with neuroticism^1–4^ ,but with opposing valence. Because the metanalytic GWAS of subjective wellbeing was composed, in large part, of continuous measures of the trait, we would not expect to observe a strong monotonic pattern of differentiation of genetic architecture between neuroticism and wellbeing across the neuroticism severity spectrum. Had the subjective well-being phenotype been an entirely all-or-nothing phenotype, with case status assigned to especially high levels of subjective well-being (i.e. the opposite tail of the severity distribution occupied by MDD), we would expect to observe the strongest absolute magnitude associations between high well-being and neuroticism at the lowest neuroticism severity levels. As expected, we observed a flat pattern of moderate-to-strong negative genetic correlations across all neuroticism severity levels (Figure S4;Table S4). A GLS analysis to did not provide evidence for either a linear ($\beta_{1}$ = -0.021, s.e. = 0.016, p = 0.176) or quadratic ($\beta_{1}$ = -0.023, s.e. = 0.017, p = 0.186;$\beta_{2}$ = 0.004, s.e. = 0.015, p = 0.782) association between neuroticism severity and the genetic association between neuroticism and subjective well-being.

***Item-level Genetic Correlations with MDD Across Item Severities***

It is possible that the differentiation of genetic architecture of negative emotionality across cut points is driven specifically by more severe neuroticism items exhibiting higher genetic correlations with MDD, rather than differentiation of genetic architecture according to severity of negative emotionality irrespective of the specific symptoms or items endorsed. Put differently, it is possible that items with higher endorsement rates (i.e. less severe items) are more genetically distinct from MDD than those with lower endorsement rates (more severe items). To test whether Neuroticism item endorsement rates were associated with their genetic correlations with MDD, we obtained item-level summary GWAS statistics of the UKB neuroticism items (<http://www.nealelab.is/uk-biobank/>). We estimated genetic correlations between each item and MDD. Following the same general pipeline applied for our primary analyses, we converted the endorsement rates for each item to a location on the normal distribution using the qnorm() function in R to compute item severities and regressed genetic correlations on these severities using GLS regression. Results are presented in Figure S5. While there was a tendency for more severe items to exhibit higher genetic correlations with MDD with a significant linear trend detected ($\beta_{1}$ = -0.262, s.e. = 0.038, P = 0.013), this association was quite weak and uneven across items, as evidenced by the considerable heterogeneity in item-MDD genetic correlations at all severity levels. These genetic correlations between neuroticism severity and MDD therefore do not appear to be primarily driven by specific neuroticism items.

#### **Re-Estimating GLS Models Using Direct Estimates of Genetic Correlation**

Our primary analyses were applied to the estimates contained within standardized genetic covariance matrix (S_Stand), and its sampling covariance matrix (V_Stand), as estimated using the ldsc() function within the Genomic SEM R package. These matrices are computed by standardizing the values within the unstandardized genetic covariance matrix (S) and its sampling covariance matrix (V) relative to the corresponding vector of heriabilities found on the S matrix’s diagonal. This transformation has the oftentimes desirable property of rendering the estimates more interpretable while maintaining the Z statistics and p values invariant across corresponding elements of S and S_Stand. Despite being nearly equivalent to a genetic correlation matrix, it may be more appropriate to refer to the S_Stand as a standardized genetic covariance matrix. The reason for this is that the sampling variances and covariances contained within V_Stand are do not correspond to the multivariate sampling distribution that would be obtained from directly estimating the genetic correlations across independent samples (or resamples of the observed sample data). This is most clear by inspecting the terms in V_Stand that correspond to the sampling variances (squared standard errors) of the standardized SNP heritabilities contained on the diagonal of S_Stand. If a genetic correlation matrix were to be estimated across independent samples, the diagonals would always be 1, and the sampling variance of these estimates would be 0. However, V_Stand reports nonzero sampling variances for these parameters, reflecting uncertainty in the SNP heritability estimates, which have simply been rescaled to the standardized metric.

The matrix of genetic correlations (R) and the corresponding matrix of their sampling covariances (V_R_) can be estimated within the Genomic SEM computational framework, by specifying a model with standardized genetic factors with fixed unit variances to underly each GWAS phenotype, setting the residual genetic variance of each GWAS phenotype to zero, and freely estimating the factor loadings (see Figure S6 for a bivariate representation of this model). Because the factors are on a standardized metric, the estimated genetic covariances are direct estimate of the genetic correlations, and the corresponding sampling variances and covariances contained within V_R_ can be interpreted as describing the expected sampling distribution of the genetic correlation estimates themselves. As would be expected for a true genetic correlation matrix, the variances on the diagonal of R are treated as fixed. We provide an R function, rgmodgen(), to estimate this model, thus estimate R and V_R_ for a given set of GWAS phenotypes.

Figure S7 plots standard errors of the estimates of the genetic correlations, drawn from the square roots of the diagonal elements of V_R_, against those of the standardized genetic covariances, drawn from the square roots of the diagonal elements of V_Stand_. We find that the SEs of the genetic correlations are generally smaller than those of the standardized genetic covariances. However, the off diagonal elements are V_R_ and V_ V_Stand_ (not represented), representing the sampling covariances, also differ. Therefore, to determine the effect of using S_Stand_ and V_Stand_ vs. R and V_R_, we conducted our primary analyses again using R and V_R_. As reported in Table S5, we find very similar patterns of results when using R and V_R in place of S_Stand_ and V_Stand_.

Supplementary Figures

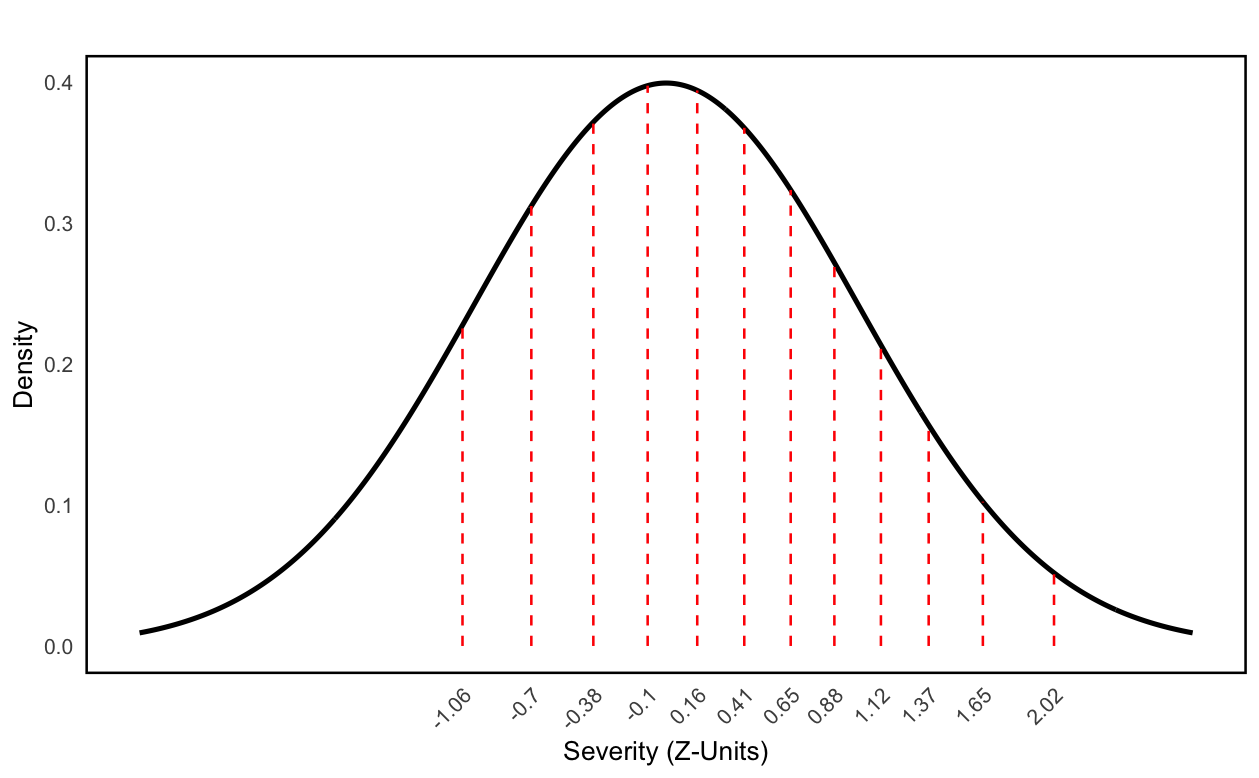

**Figure S1** Mapping of neuroticism cut points to the theoretical normal distribution. Each cut point is represented by a single line that has been mapped onto the theoretical normal distribution based on the percentage of individuals above that cut point (using the qnorm() function in R).

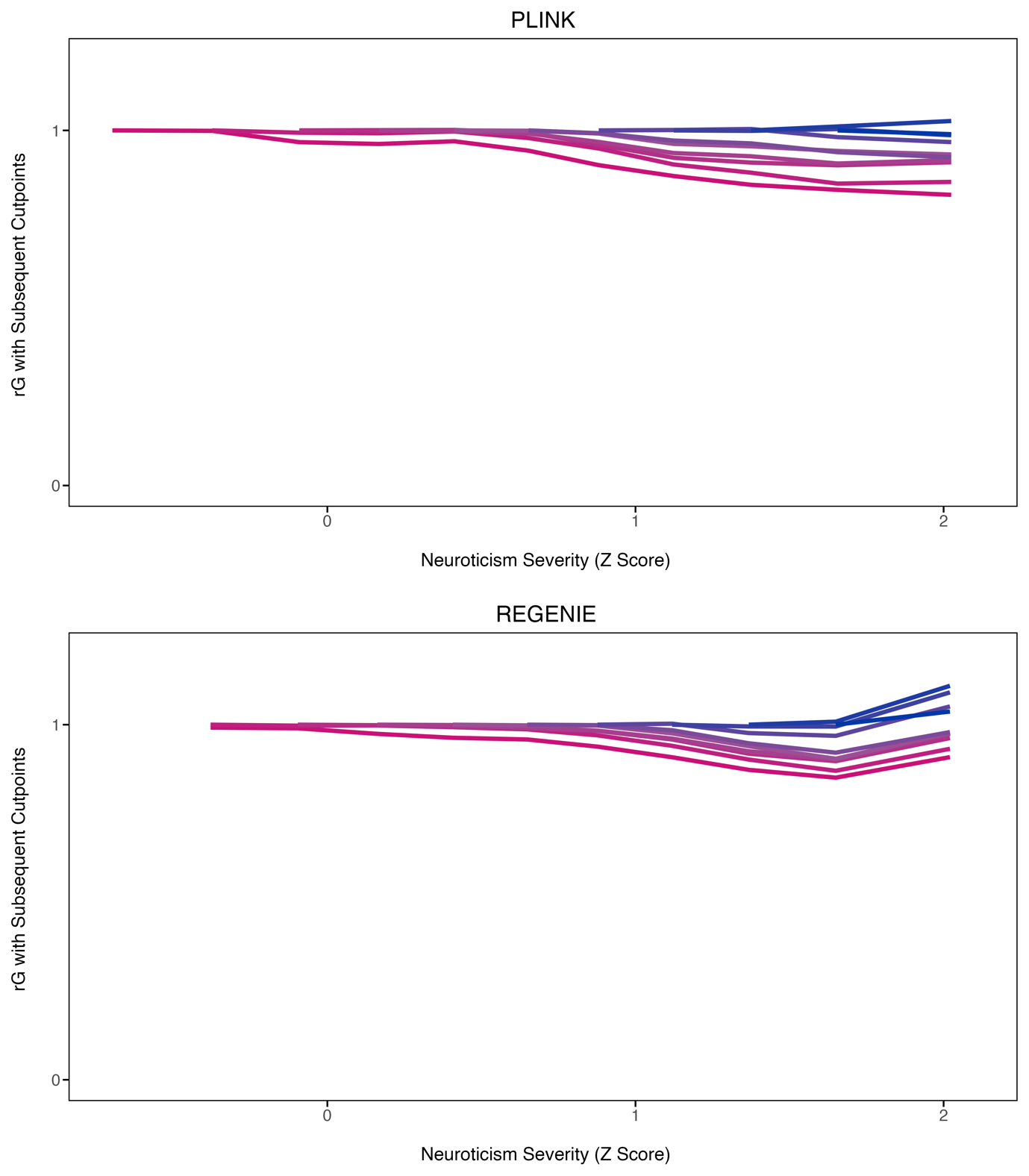

**Figure S2** Genetic correlations between neuroticism cut points. Top panel uses GWAS of neuroticism in N= 414,448 individuals using REGENIE to correct for relatedness. Bottom panel uses genome wide association study (GWAS) of Neuroticism in N = 281,993 unrelated individuals using PLINK. Each line follows the genetic correlations from the point at the start of the line with the subsequent cut points. Genetic correlations were estimated by applying linkage disequilibrium score regression (LDSC) to the GWAS summary statistics.

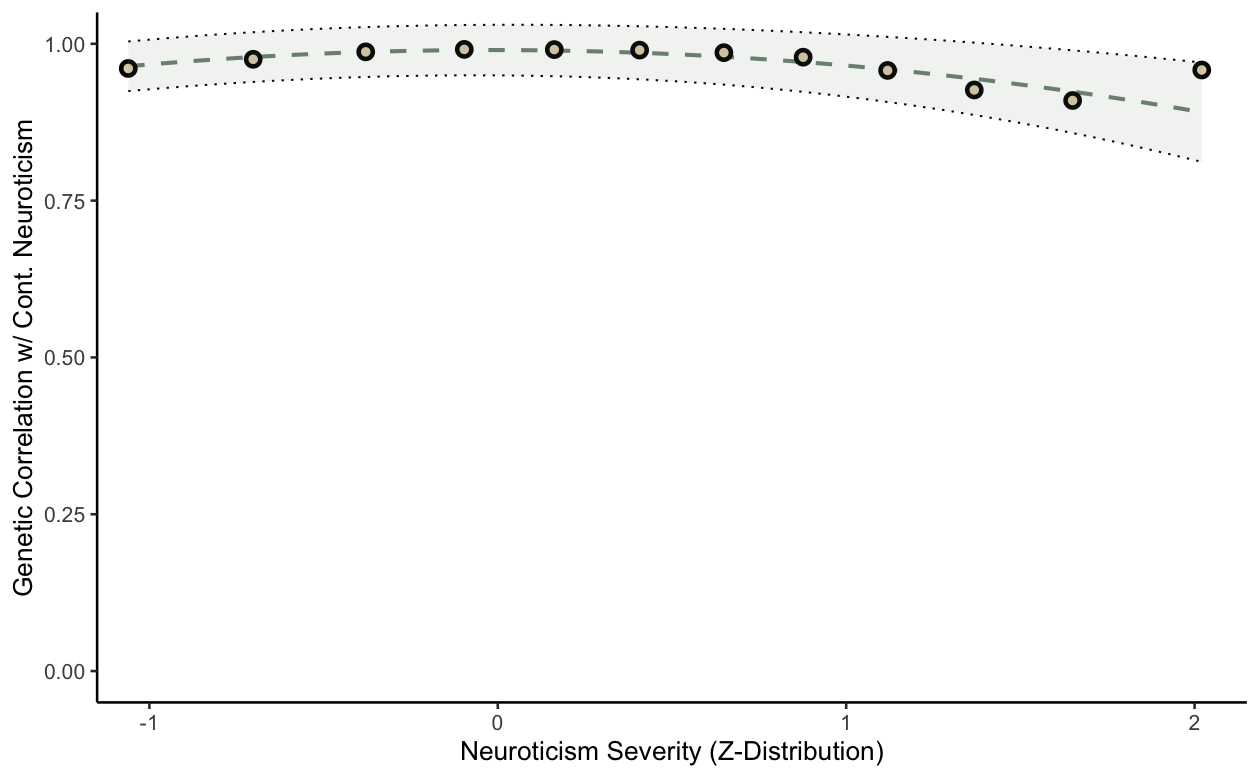

**Figure S3** Pattern of genetic correlations between neuroticism cut points, and a continuous measure of neuroticism. With the horizontal axis showing the proportion of individuals below the given cut point mapped onto the theoretical normal distribution using the qnorm() function in R. The line of best fit is shown, based on a quadratic regression model, with parameters estimated using generalized least squares. Shading around the line of best fit represents +/- 1 SE. GLS analyses indicated a non-significant linear ($\beta_{1}$ = -0.01, s.e. = 0.01, P = 0.365).), but significant quadratic trend ($\beta_{1}$ = 0.001, s.e. = 0.01, P = 0.934; $\beta_{2}$ = -0.03, s.e. = 0.01, P = 0.0013), with the highest genetic correlations being found between the more central cut points near the mean neuroticism score in the sample in which the summary statistics were derived.

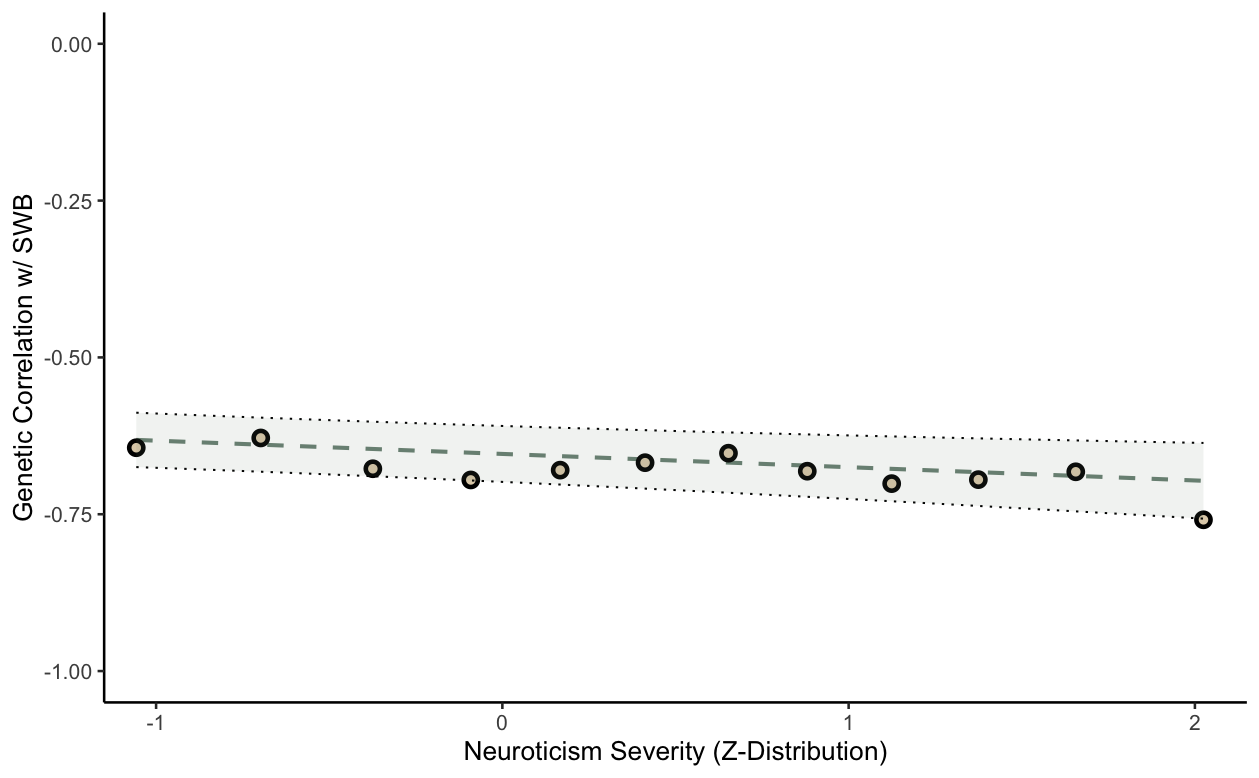

**Figure S4** Genetic correlations between a meta-analysis of subjective wellbeing (SWB) and each neuroticism cut point. The line of best fit is shown, based on a linear regression model, with parameters estimated using generalized least squares (GLS). Shading around the line of best fit represents +/- 1 s.e.. , The results of the GLS analysis estimated both the linear ($\beta_{1}$ = -0.021, s.e. = 0.02, P = 0.186), and quadratic ($\beta_{1}$ = -0.023, s.e. = 0.02, P = 0.186; $\beta_{2}$ = 0.004, s.e. = 0.01, P = 0.783) terms in the regression equation to be non-significant

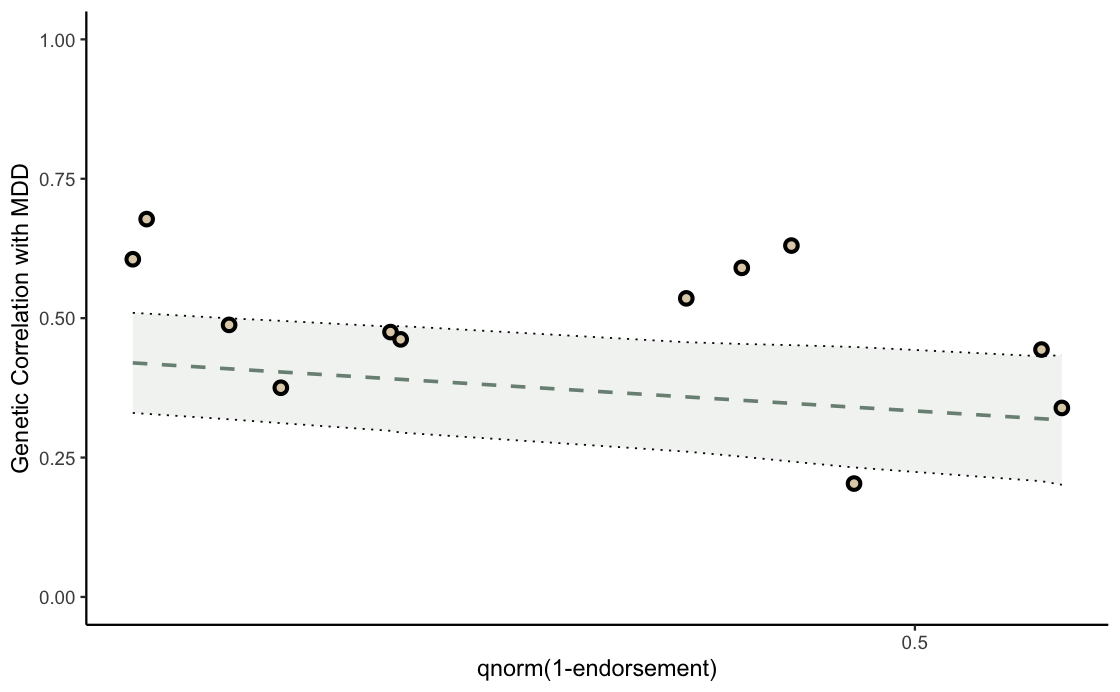

**Figure S5** Genetic correlations between each individual neuroticism item (summary statistics for the GWAS of each neuroticism item acquired from <http://www.nealelab.is/uk-biobank/>) and major depressive disorder (MDD). The horizontal axis represents the proportion of individuals endorsing a given item mapped to the theoretical normal distribution with the qnorm() function in R. The line of best fit is based on a linear regression model, with parameters estimated using Generalized Least Squares (GLS). Shading around the line represents +/- 1 SE. GLS analyses indicated a significant linear ($\beta_{1}$ = -0.262, s.e. = 0.038, P = 0.013) but not quadratic trend ($\beta_{1}$ = -0.162, P = 0.039 $\beta_{2}$ = 0.176, s.e. = 0.122, p = 0.085). However, this association was quite weak, as evidenced by the considerable heterogeneity in item-MDD genetic correlations at all severity levels.

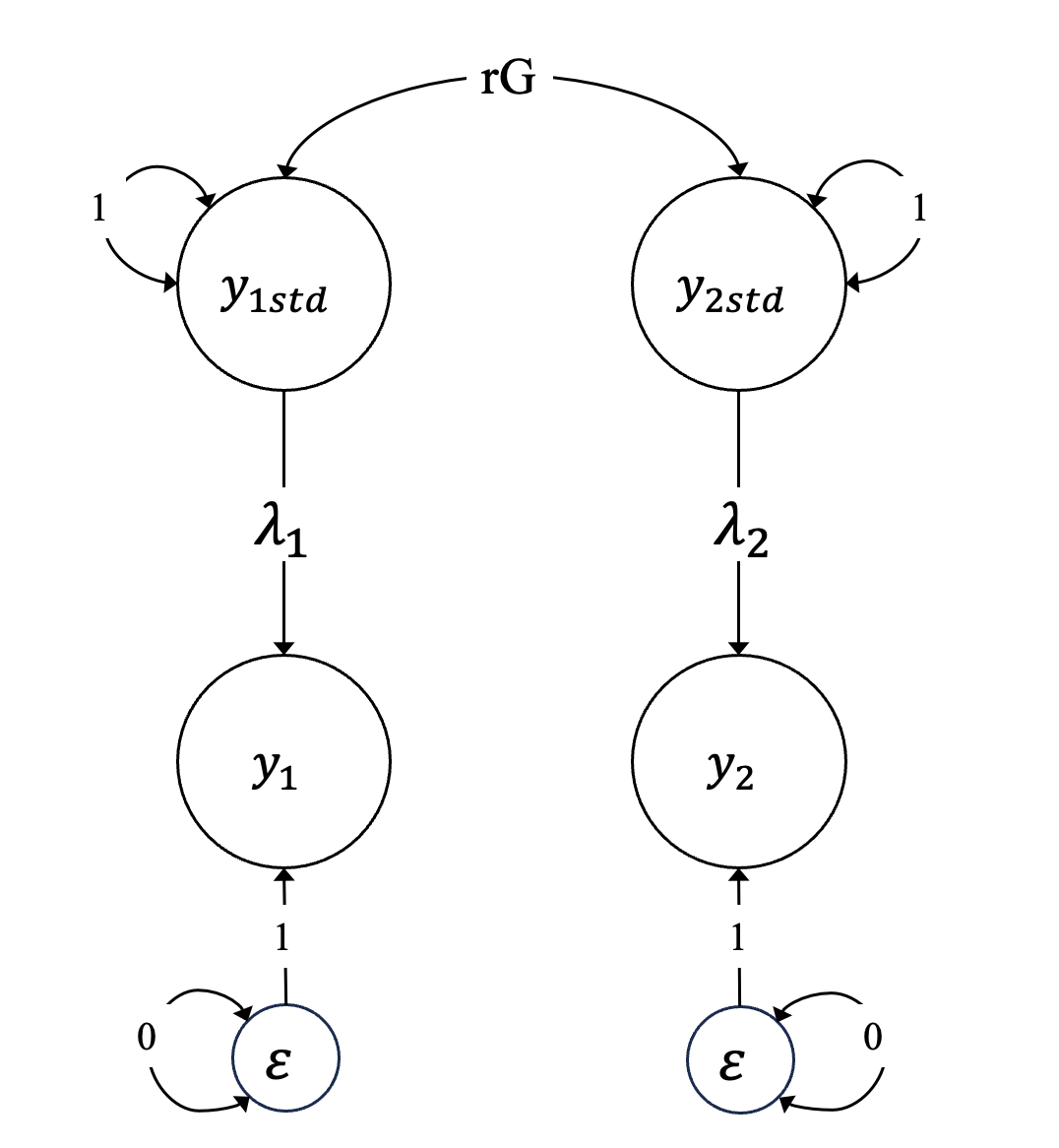

**Figure S6.** A path diagram representing the bivariate version of the Genomic SEM model that is used to obtain direct estimates of genetic correlations (R) and its sampling covariance Matrix (V_R_). Standardized genetic factors are specified to underly each GWAS phenotype; the residual genetic variance of each GWAS phenotype is to zero, and each factor loading is freely estimated (yielding an estimate of the square root of the SNP heritability of the corresponding GWAS phenotype). Because the factors are on a standardized metric, the estimated genetic covariance is a direct estimate of the genetic correlation, and the corresponding SE can be interpreted as the expected standard deviation of the sampling distribution of that genetic correlation estimate.

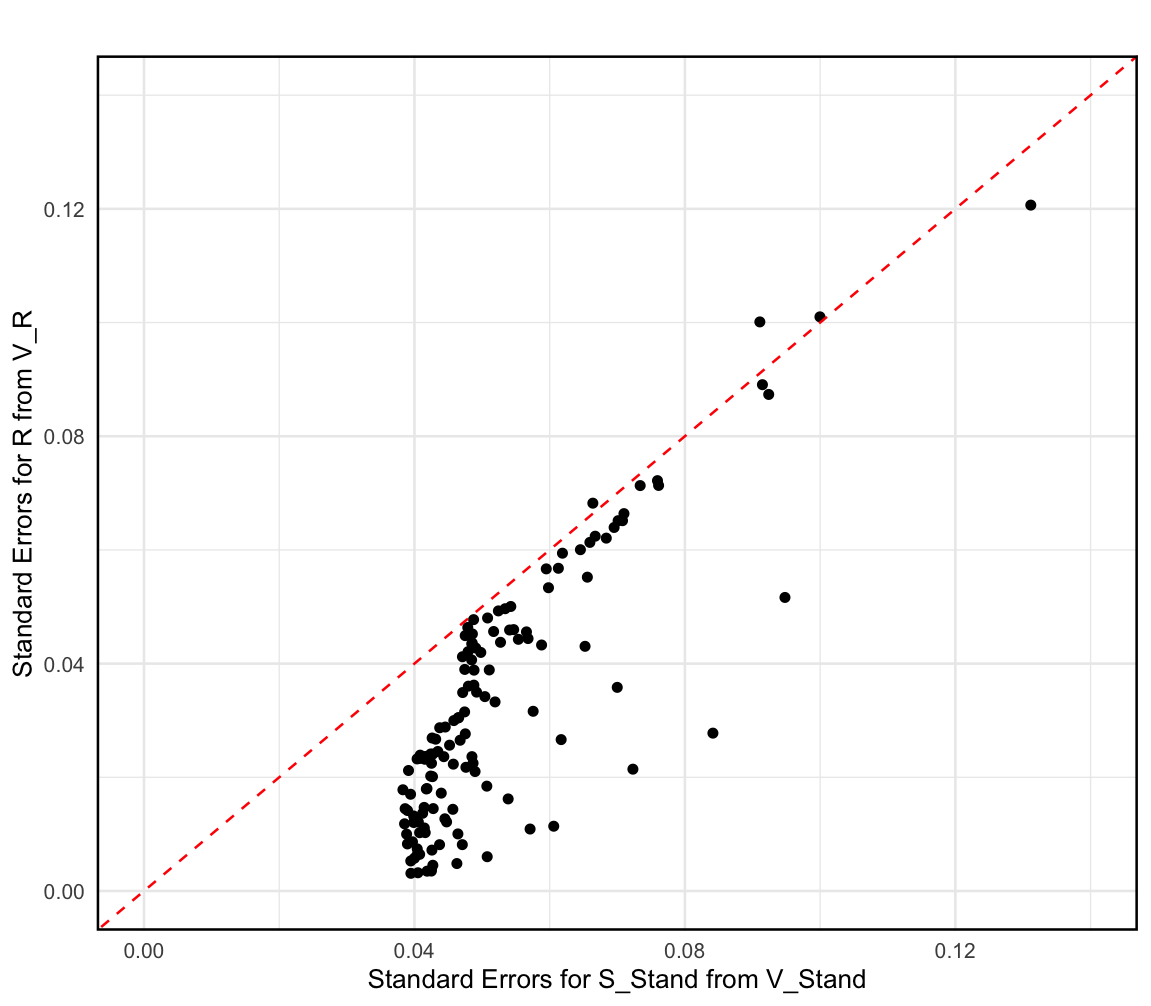

**Figure S7** Comparison of the standard errors of the estimates of genetic correlations from our primary analyses, drawn from the square roots of the diagonals of V_Stand_ and V_R_ matrices. Red line at x = y. The squared standard errors found on the diagonal of V_Stand_ are the rescaled values from the V matrix that maintain the p values and Z statistics of the genetic covariances, whereas those found on the diagonal of V_R_ are estimated using a multivariate version of the model illustrated in Figure S6.

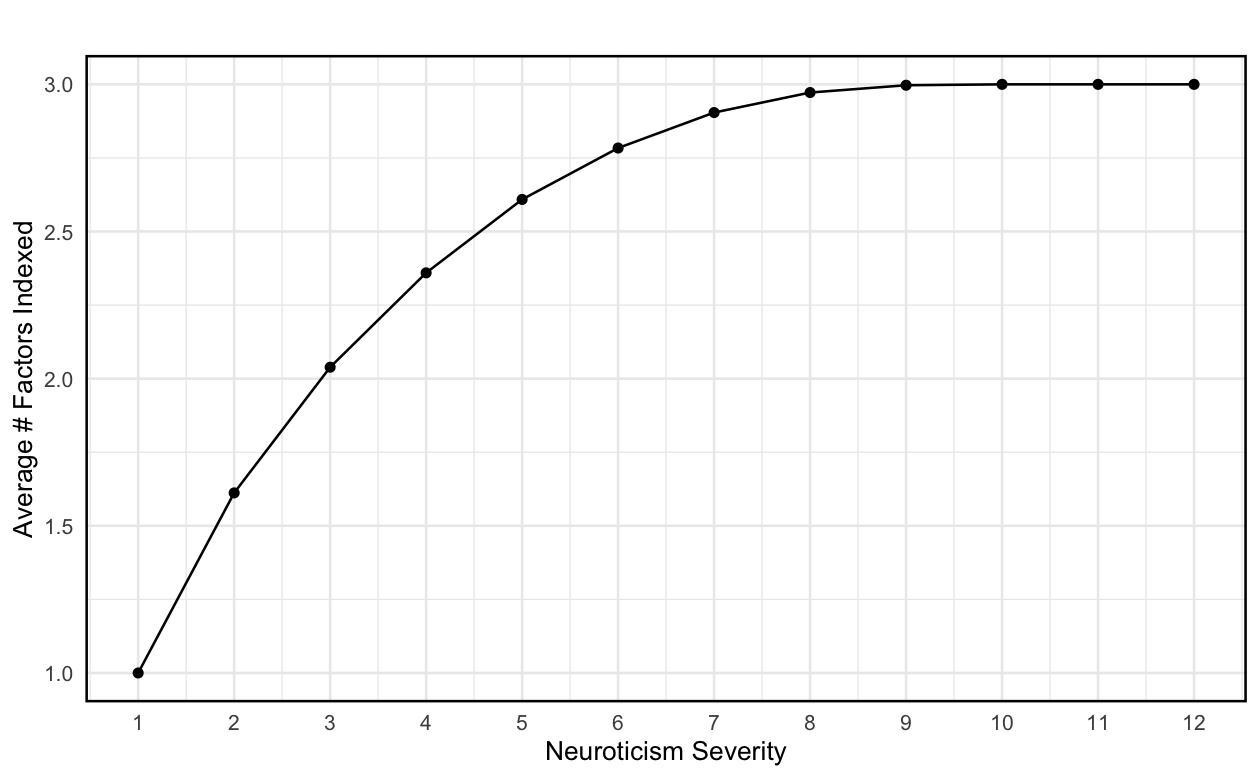

**Figure S8** Average number of neuroticism facets indexed by neuroticism severity. We used the three-factor model of neuroticism from Grotzinger et al., 2019^5^ to compute the average number of sub-factors endorsed with at least one item at each level of severity.

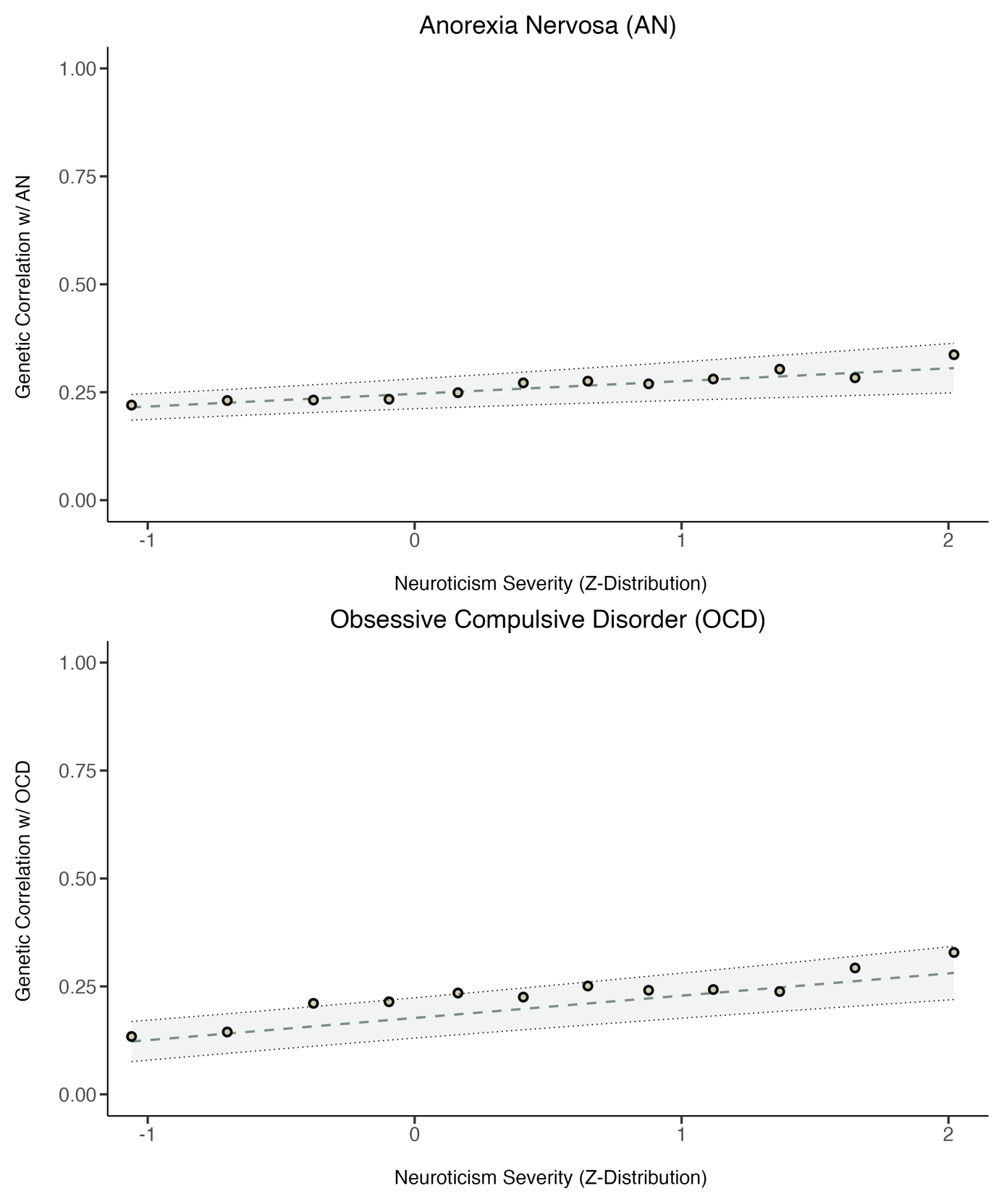

**Figure S9** Genetic correlations between neuroticism and each of two compulsive disorders (Anorexia Nervosa, Obsessive Compulsive Disorder (OCD)) as a function to neuroticism severity. The line of best fit is shown with parameters estimated using Generalized Least Squares (GLS) regression. GLS regression indicated that there were significant linear trends in the genetic correlations between neuroticism severity, and OCD and Anorexia. GLS estimates are reported in Table S7.

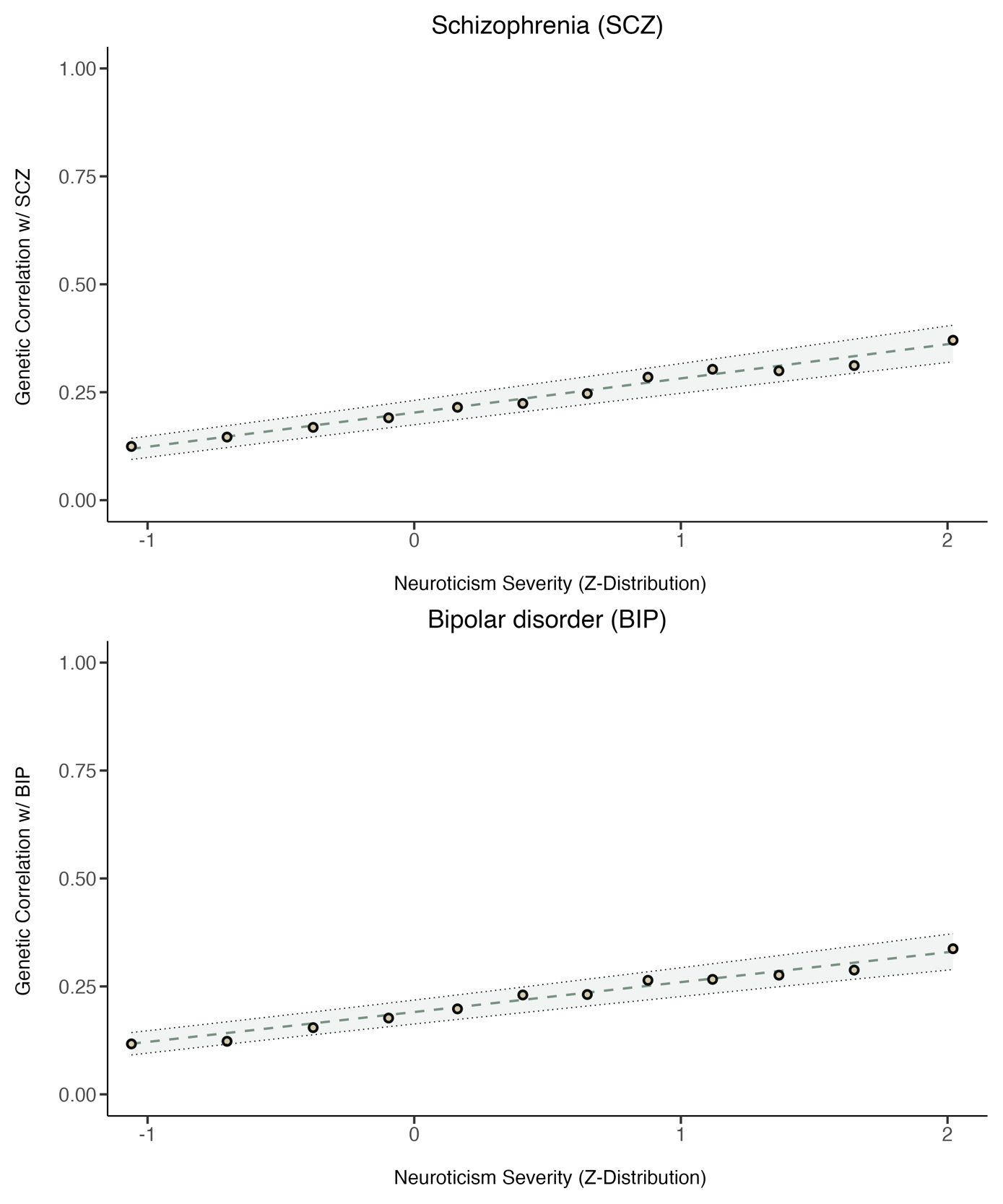

**Figure S10** Genetic correlations between neuroticism and each of two thought disorders (Schizophrenia (SCZ), and Bipolar Disorder (BIP)) as a function of neuroticism severity. The line of best fit is shown with parameters estimated using Generalized Least Squares (GLS) regression. GLS regression indicated that there were significant linear trends in the genetic correlations between neuroticism severity and both SCZ and BIP. GLS estimates are reported in Table S7.

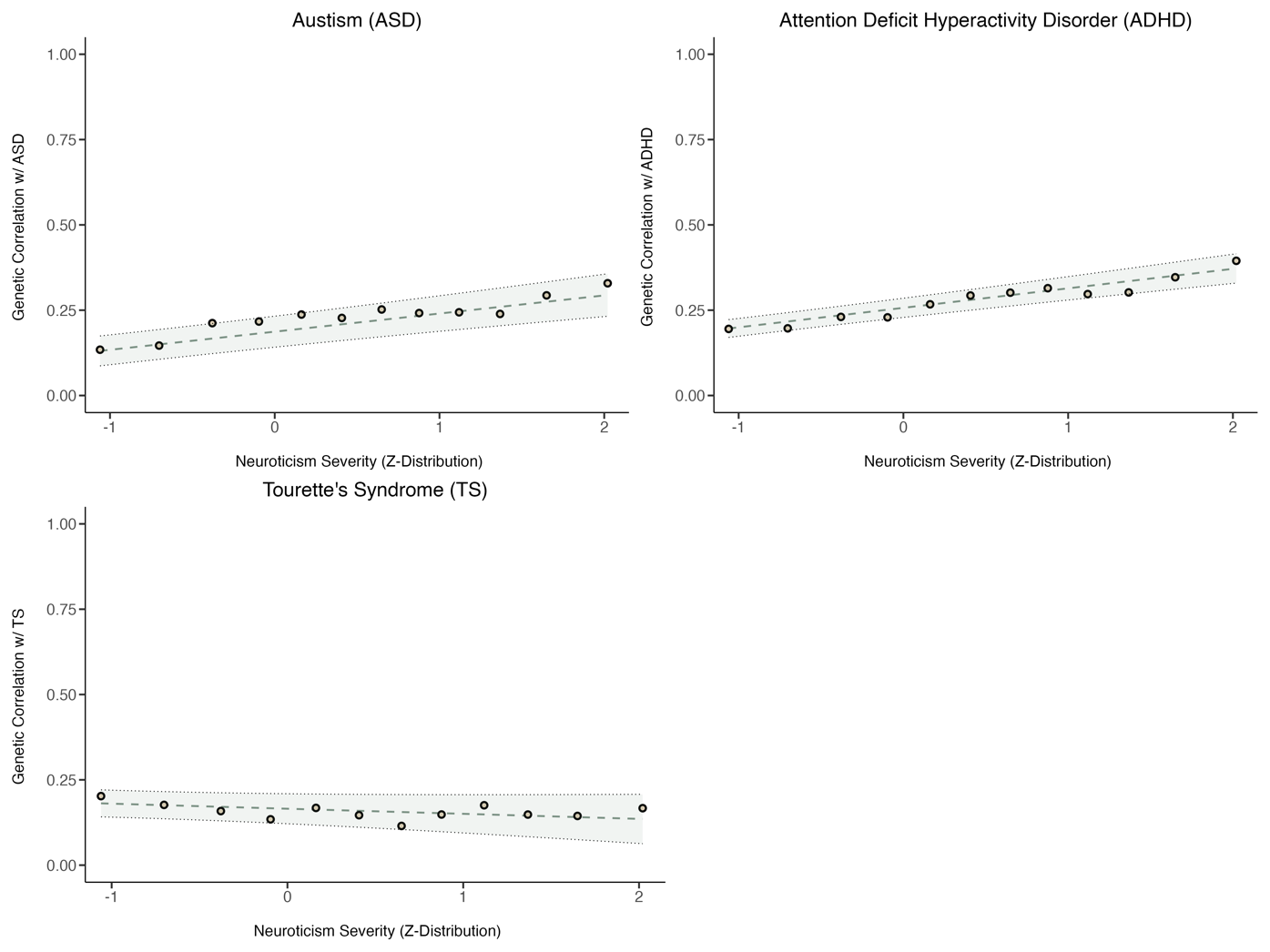

**Figure S11** Genetic correlations between neuroticism and each of two neurodevelopmental disorders (Autism, and Attention Deficit Hyperactivity Disorder (ADHD), Tourette’s Syndrome (TS)) as a function of neuroticism severity. The line of best fit is shown with parameters estimated using Generalized Least Squares (GLS) regression. GLS regression indicated that there were significant linear trends in the genetic correlations between neuroticism severity, and both Autism and ADHD, but not TS. GLS estimates are reported in Table S7.

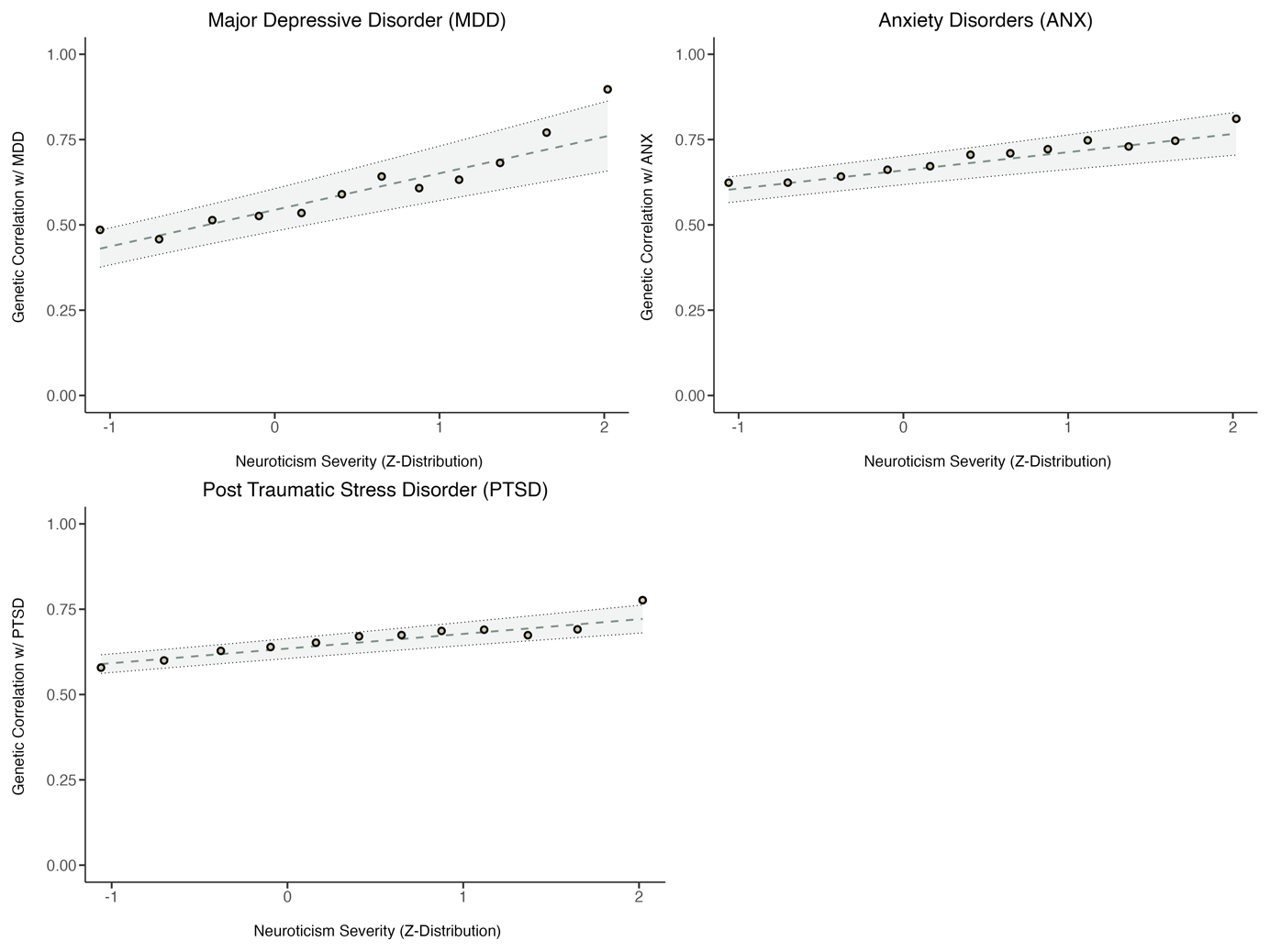

**Figure S12** Genetic correlations between neuroticism and each three internalizing disorders (Post Traumatic Stress Disorder (PTSD), Anxiety Disorders (ANX), and Major Depressive Disorder (MDD)) as a function of neuroticism severity. The line of best fit is shown with parameters estimated using Generalized Least Squares (GLS) regression. GLS regression indicated that there were significant linear trends in the genetic correlations between neuroticism severity, and MDD and PTSD, but not ANX, possibly due to insufficient power. GLS estimates are reported in Table S7.

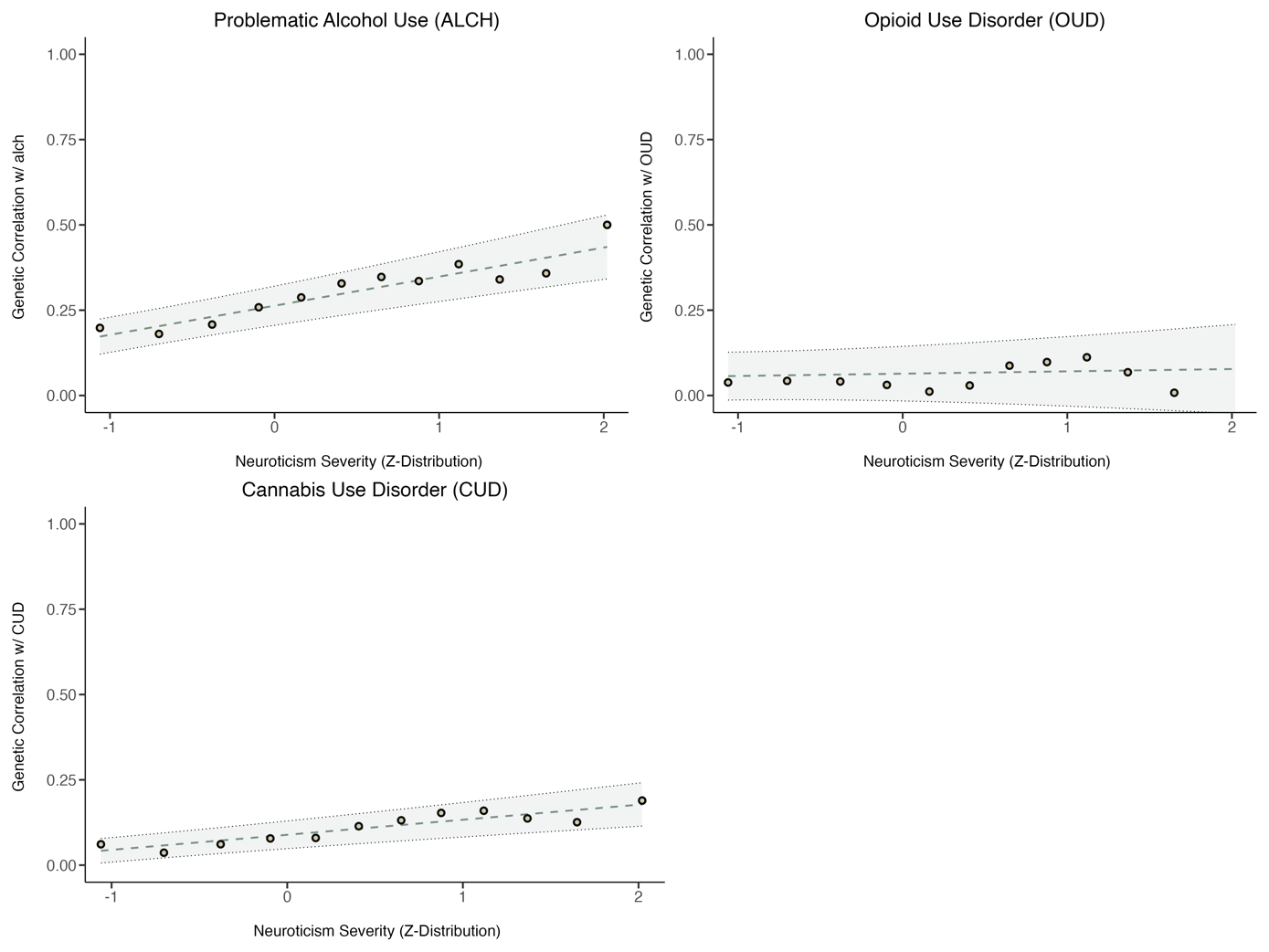

**Figure S13** Genetic correlations between neuroticism and each of three substance use disorders (Problematic Alcohol Use (ALCH), Cannabis Use Disorder (CUD), and Opioid Use Disorder (OUD)) as a function of neuroticism severity. The line of best fit is shown with parameters estimated using Generalized Least Squares (GLS) regression. GLS regression indicated that there were significant linear trends in the genetic correlations between neuroticism severity, and ALCH and CUD, but not OUD. GLS estimates are reported in Table S7.

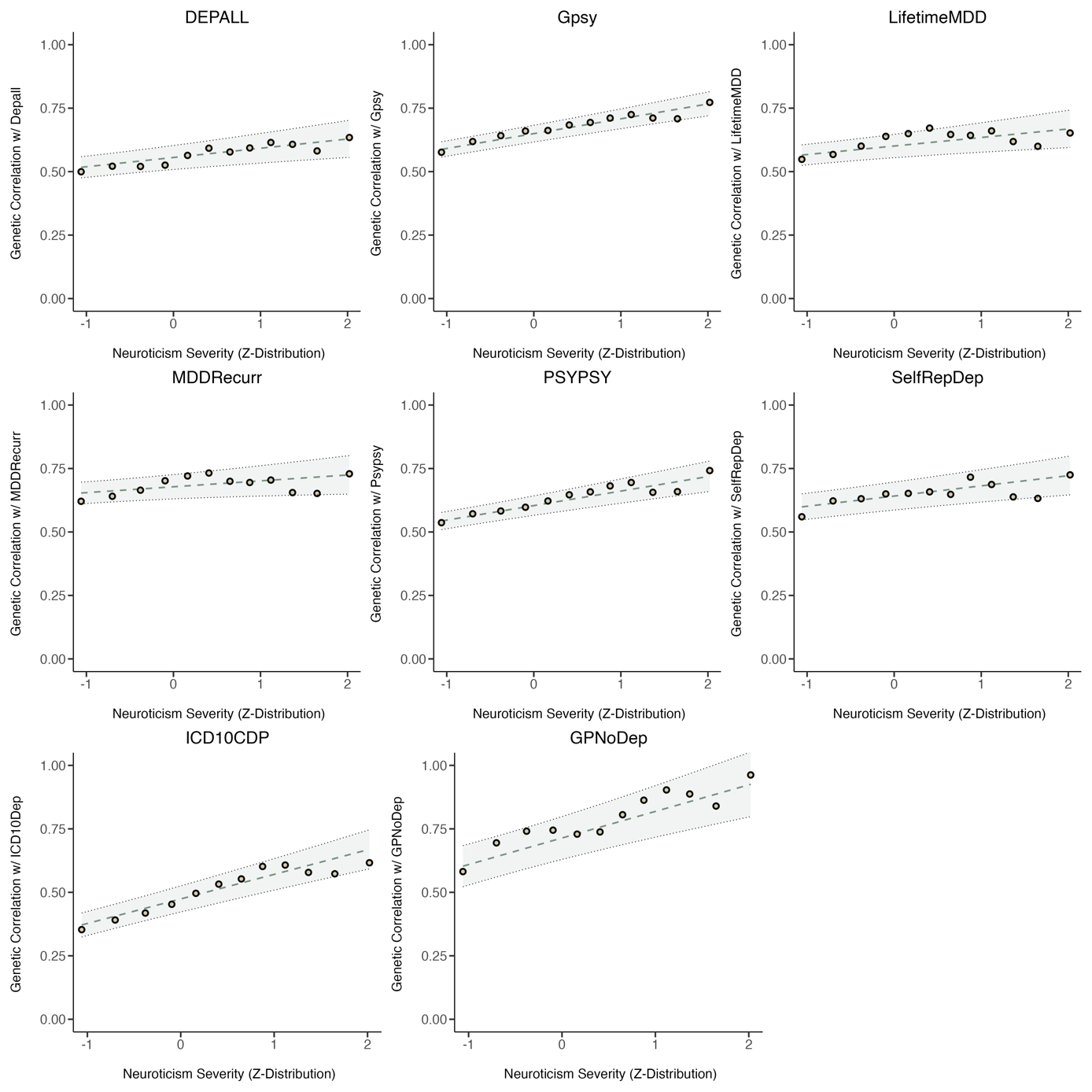

**Figure S14.** Genetic correlations between neuroticism and each of the eight binary depression phenotypes from Cai et al., (2020)^6^ as a function of neuroticism severity. The line of best fit is shown with parameters estimated using Generalized Least Squares (GLS) regression. GLS regression indicated that there were significant linear trends in the genetic correlations between neuroticism severity, and all GWAS phenotypes except for LifetimeMDD and MDDRecurr. GLS estimates are reported in Table S9.
